# Comparative single-dose mRNA and ChAdOx1 vaccine effectiveness against SARS-CoV-2, including early variants of concern: a test-negative design, British Columbia, Canada

**DOI:** 10.1101/2021.09.20.21263875

**Authors:** Danuta M Skowronski, Solmaz Setayeshgar, Macy Zou, Natalie Prystajecky, John R Tyson, Hind Sbihi, Chris D Fjell, Eleni Galanis, Monika Naus, David M Patrick, Shiraz El Adam, May Ahmed, Shinhye Kim, Bonnie Henry, Linda M N Hoang, Manish Sadarangani, Agatha N Jassem, Mel Krajden

**Author notes:** **Corresponding Author:** Danuta M Skowronski MD, FRCPC, 655 West 12th Avenue, Vancouver, British Columbia, Canada V5Z 4R4. **Alternate corresponding author:** Solmaz Setayeshgar PhD, 655 West 12th Avenue, Vancouver, British Columbia, Canada V5Z 4R4.

## Abstract

**Introduction:** In randomized controlled trials, single-dose efficacy against SARS-CoV-2 illness exceeded 90% for mRNA vaccines (BNT162b2 and mRNA-1273), and 75% for ChAdOx1. In British Columbia (BC), Canada second doses were deferred up to 16 weeks and ChAdOx1 was only initially recommended for adults 55 years of age and older. We compared single-dose vaccine effectiveness (VE) during the spring 2021 wave in BC when Alpha and Gamma variants of concern (VOC) predominated.

**Methods:** VE was estimated against infection and hospitalization by test-negative design: cases were RT-PCR test-positive for SARS-CoV-2 and controls were test-negative. Adults 50-69 years old with specimen collection between April 4 and May 22 (weeks 14-20) were included. Variant-specific VE was estimated between weeks 17-20 when genetic characterization of all case viruses was performed, primarily through whole genome sequencing.

**Results:** VE analyses included 7,116 (10%) cases and 60,958 controls. Three-quarters of vaccinated participants received mRNA vaccine (60% BNT162b2, 15% mRNA-1273) and 25% received ChAdOx1. Half of genetically characterized viruses were Alpha, with 38% Gamma, 4% Delta and 8% non-VOCs. Single-dose VE against any infection was 75% (95%CI: 72-78) for BNT162b2, 82% (95%CI: 76-87) for mRNA-1273 and 61% (95%CI: 54-66) for ChAdOx1. VE against hospitalization was 83% (95%CI: 76-89), 85% (95%CI: 63-94) and 96% (95%CI: 86-99), respectively. VE against Alpha vs. Gamma infections did not differ among mRNA (78%;95%CI: 73-82 and 80%;95%CI: 74-85) or ChAdOx1 (66%;95%CI: 57-74 and 60%;95%CI: 48-69) recipients.

**Conclusions:** A single dose of mRNA vaccine reduced the SARS-CoV-2 infection risk by at least 75%, including infections due to early VOC. Although effectiveness of a single dose of ChAdOx1 was lower at 60% against infection, just one dose of any vaccine reduced the hospitalization risk by more than 80%. In the context of constrained vaccine supplies, these findings have implications for global vaccine deployment to reduce the overall burden of infections and hospitalizations due to SARS-CoV-2.

## INTRODUCTION

The first SARS-CoV-2 vaccine to be authorized in Canada was an mRNA formulation, BNT162b2 (Pfizer-BioNTech), approved on December 9, 2020 for individuals ≥16-years of age at a schedule of two doses spaced 3 weeks apart [1]. Thereafter, mRNA-1273 (Moderna), was authorized on December 23 for those ≥18 years old as two doses spaced 4 weeks apart. Chimpanzee adenoviral vectored (ChAdOx1) vaccine (AstraZeneca and equivalent COVISHIELD), was authorized on February 26, 2021 for ≥18-years-olds as two doses spaced 4-12 weeks apart. In randomized-controlled trial (RCT) analyses, single-dose efficacy against SARS-CoV-2 illness was 92-93% for mRNA vaccines [2,3], and 76% for ChAdOx1 [4]. In the context of constrained vaccine supplies and heightened epidemic activity, Canada’s National Advisory Committee on Immunization (NACI) recommended on March 3, 2021 (epidemiological week 9) that the interval between first and second doses of all SARS-CoV-2 vaccines be extended up to 16 weeks to enable more people to benefit as rapidly as possible from substantial single-dose protection [5]. On March 29 (week 13), NACI also recommended that ChAdOx1 be limited to adults ≥55-years-old due to emerging global reports of vaccine-associated thrombosis with thrombocytopenia, with that age-restriction subsequently lowered to ≥30 years on April 23 (week 16) [6,7].

Community vaccination in British Columbia (BC), Canada followed a sequential age-based strategy, prioritizing mRNA vaccines for the eldest citizens beginning around week 10. The spring 2021 pandemic wave in BC was comprised of unique co-dominance by Alpha and Gamma variants of concern (VOC), with lesser Delta contribution [8-10]. Using a test-negative design (TND) we previously reported single-dose mRNA (foremost BNT162b2) vaccine effectiveness (VE) among adults ≥70-years-old, showing the infection risk was reduced by about two-thirds overall and comparably against Alpha and Gamma variants [11]. Too few older adults had received ChAdOx1 to enable its VE estimation, but this became feasible when adults 50-69-years-old became vaccine-eligible around week 14. Herein we apply the same TND approach to compare single-dose mRNA and ChAdOx1 VE against SARS-CoV-2 infection and hospitalization, including due to VOC, among adults 50-69-years-old in BC, Canada.

## METHODS

### Source population

There are ∼1.4 million adults 50-69-years-old in BC (27% of the 5.2 million population) of whom 51% are 50-59 years and 51% are women [12]. A publicly-funded, mostly symptom-based approach for PCR-based SARS-CoV-2 diagnostic testing is broadly accessible in BC. The spring 2021 pandemic wave peaked overall in BC in week 13 with 12% of specimens testing positive and later among 50-69-year-olds in week 16 with 12% testing positive [8]. The peak-week tally of cases was 8000 in week 14, of which ∼20% were adults 50-69-years-old [8]. About 60% of the BC population resides in the Greater Vancouver Area (GVA)[11], comprised of Vancouver Coastal and Fraser Health Authorities (VCHA and FHA), the latter most affected by the spring 2021 wave [8]. With limits on travel abroad and other control measures [13,14], the epidemic gradually subsided, although test-positivity continued to exceed 5% until about week 21[8].

### Study design and analysis period

We estimated single-dose VE by TND, using multivariable logistic regression to derive the adjusted odds ratio (AOR) for vaccination among SARS-CoV-2 test-positive cases versus test-negative controls. VE and 95% confidence intervals (CI) were computed as (1-AOR) x100%. Adjusted models included age, gender, epidemiological week and health authority (HA).

The full analysis period included specimens collected from April 4-May 22 (weeks 14-20) whereas variant-specific analysis spanned April 25-May 22 (weeks 17-20). The latter was chosen balancing the capacity to genetically characterize all contributing case viruses, and the vaccine coverage and infection rates required for a sufficient number of vaccinated, variant-specific cases driving statistical power.

### Case and control selection

Cases included any infection, as well as hospitalization on or within 30 days after specimen collection. Individuals could contribute a single test-positive specimen as cases and were censored thereafter. In variant-specific analyses, cases were categorized as due to VOC (Alpha, Beta, Gamma, Delta) or non-VOC, the latter also including variants of interest (VOI) in primary analysis but excluding them in sensitivity analyses. Detailed methods for genetic characterization are provided in **Supplementary Material 1**, predicated foremost on whole genome sequencing but also RT-PCR single-nucleotide polymorphism screen for Alpha and Gamma variants [10,15,16].

Three approaches were explored for test-negative control selection: (1) all negative specimens; (2) only the most recent negative specimen; or (3) only one randomly-selected negative specimen.

### Vaccine status definition

Clients with record of a single dose of mRNA or ChAdOx1 vaccine on or before the date of specimen collection were considered vaccinated; those without such record were considered unvaccinated. Among adults 50-69-years-old with both dates available, the mean/median interval between onset and respiratory specimen collection date was 3/2 days, respectively, with interquartile range of 1-4 days. We based primary VE analyses on vaccine status defined by receipt ≥21 days before specimen collection, but assessed a range of intervals.

### Data sources and exclusions

Specimens were sampled from the provincial database that captures all RT-PCR testing for SARS-CoV-2 in BC along with client, collection and testing details although symptoms and onset dates are not consistently captured. Hospitalized cases were identified through linkage with the notifiable disease list. Vaccination information was obtained from the provincial immunization registry which captures all SARS-CoV-2 vaccinations in BC along with client and vaccination details. Individual-level database linkages were achieved through unique personal identifiers.

Specimens with invalid or missing linkage, exposure, outcome or covariate information were excluded as were specimens collected from individuals: identified as cases before the analysis period; vaccinated more than once; residents of long-term care, assisted-living or independent-living facilities; or tested outside of public funding owing to systematically lower likelihood of test-positivity [8]. This also excludes individuals routinely screened as part of travel requirements abroad.

### Ethics statement

Data linkages and analyses were authorized by the Provincial Health Officer under the Public Health Act and exempt from research ethics board review.

## RESULTS

### Case and control profiles

In total, 68,074 SARS-CoV-2 specimens contributed to VE analyses, including 7116 (10%) test-positive cases and 60,958 test-negative controls (**Supplementary Figure 2**). Decrease in test-positivity and case numbers by successive week mirrored provincial surveillance patterns (**Table 1**)[8]. Case and control distributions reflected population distributions by age and gender, with variation by HA reflecting regional differences in epidemic intensity [8,12].

**Table 1.** Participant characteristics by case and control status, adults 50-69 years of age, British Columbia (BC), Canada, weeks 14-20

| Characteristic | Overall Participants |  | Test-positive Cases Any infection |  | Test-positive Cases Hospitalized<sup>a</sup> |  | Test-negative Controls<sup>b</sup> |  |
| --- | --- | --- | --- | --- | --- | --- | --- | --- |
|  | n | %<sup>c</sup> | n | %<sup>c</sup> | n | %<sup>c</sup> | n | %<sup>c</sup> |
| **Overall 50-69 years N (%)** | 68 074 | NA | 7116 | 10.5<sup>d</sup> | 716 | 10.1<sup>e</sup> | 60 958 | 89.5<sup>f</sup> |
| **Age group (years)** |  |  |  |  |  |  |  |  |
| 50-59 | 38 730 | 56.9 | 4242 | 59.6 | 327 | 45.7 | 34 488 | 56.6 |
| 60-69 | 29 344 | 43.1 | 2874 | 40.4 | 389 | 54.3 | 26 470 | 43.4 |
| Median age (Interquartile range) | 58 | 54-63 | 58 | 53-63 | 60 | 55-65 | 58 | 54-63 |
| **Gender** |  |  |  |  |  |  |  |  |
| Male | 32 096 | 47.0 | 3650 | 51.3 | 427 | 59.6 | 28 446 | 46.7 |
| Female | 35 978 | 53.0 | 3466 | 48.7 | 289 | 40.4 | 32 512 | 53.3 |
| **Epidemiological week** |  |  |  |  |  |  |  |  |
| 14 | 12 678 | 18.6 | 1530 | 21.5 | 131 | 18.3 | 11 148 | 18.3 |
| 15 | 11 833 | 17.4 | 1420 | 20.0 | 155 | 21.7 | 10 413 | 17.1 |
| 16 | 10 238 | 15.0 | 1207 | 17.0 | 109 | 15.2 | 9031 | 14.8 |
| 17 | 10 303 | 15.1 | 1076 | 15.1 | 126 | 17.6 | 9227 | 15.1 |
| 18 | 9068 | 13.3 | 803 | 11.3 | 74 | 10.3 | 8265 | 13.6 |
| 19 | 7539 | 11.1 | 666 | 9.4 | 83 | 11.6 | 6873 | 11.3 |
| 20 | 6415 | 9.4 | 414 | 5.8 | 38 | 5.3 | 6001 | 9.8 |
| **Health authority** |  |  |  |  |  |  |  |  |
| Fraser (FHA) | 34 807 | 51.1 | 4118 | 57.9 | 403 | 56.3 | 30 689 | 50.3 |
| Interior (IHA) | 9511 | 14.0 | 685 | 9.6 | 59 | 8.2 | 8826 | 14.5 |
| Northern (NHA) | 1935 | 2.8 | 292 | 4.1 | 30 | 4.2 | 1643 | 2.7 |
| Vancouver Coastal (VCHA) | 14 563 | 21.4 | 1745 | 24.5 | 208 | 29.1 | 12 818 | 21.0 |
| Vancouver Island (VIHA) | 7258 | 10.7 | 276 | 3.9 | 16 | 2.2 | 6982 | 11.5 |

<sup>a</sup> Excludes hospitalizations prior to or more than 30 days after specimen collection<sup>b</sup> As per Approach 1 for control selection: includes all test-negative specimens collected from individuals before the end of the analysis period or becoming a test-positive case.<sup>c</sup> Unless otherwise specified, percentage reflects distribution by characteristic (i.e. column percentage)<sup>d</sup> Percentage displayed is percentage of participants overall that are cases<sup>e</sup> Percentage displayed is percentage of cases hospitalized<sup>f</sup> Percentage displayed is percentage of participants overall that are controls

### Vaccination profiles

The percentage vaccinated among controls was similar to the provincial vaccine coverage among 50-69-year-olds by epidemiological week 14 (16% vs 18%), 15 (24%% vs. 29%), 16 (35% vs. 39%), 17 (46% vs. 50%), 18 (58% vs. 62%), 19 (68% vs. 72%) and 20 (75% vs. 76%) (**Table 2**). The longest follow-up period post-vaccination was 111 days for cases and 143 days for controls; however, >80% of specimens were collected <42 days since first vaccination. Three-quarters of vaccinated individuals had received an mRNA vaccine: 60% BNT162b2 and 15% mRNA-1273. ChAdOx1 vs. mRNA recipients were slightly younger (median 58 vs. 61 years), and less often women (50% vs. 57%) (**Supplementary Table 2**). More ChAdOx1 than mRNA recipients resided in the GVA (89% vs. 75%), notably FHA (68% vs. 53%).

**Table 2.**
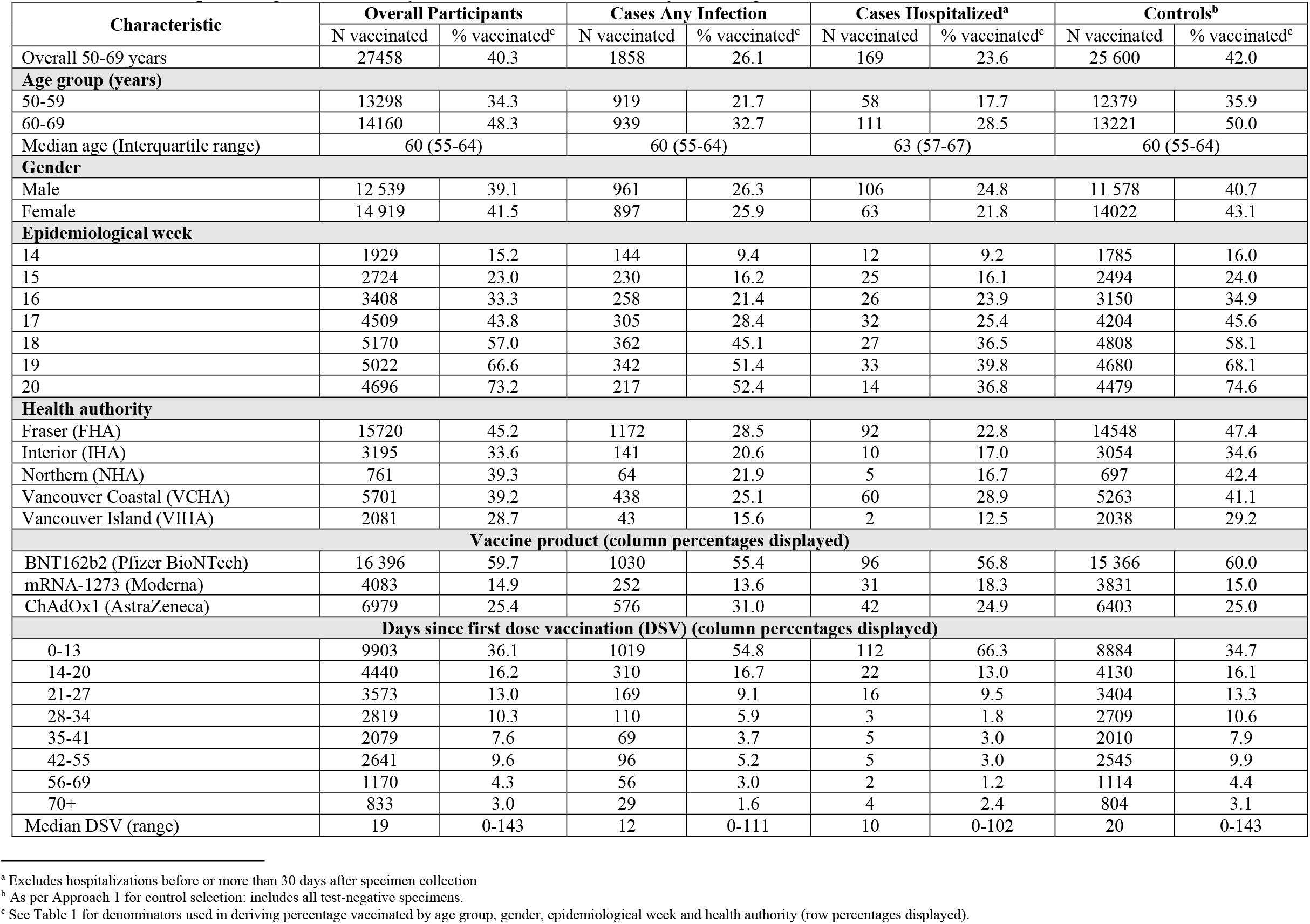
Number and percentage vaccinated by characteristic, adults 50-69 years of age, British Columbia (BC), Canada, weeks 14-20

### VOC profiles

Between weeks 17-20, 2956 cases contributed to VE analyses (**Table 1**). Of these, 2798 (95%) had sufficient specimen for genetic characterization: 2424 (87%) by whole genome sequencing and 374 (13%) by RT-PCR single-nucleotide polymorphism screen (260 Alpha and 114 Gamma) (**Supplementary Table 1**). After excluding 141 (5%) characterized case viruses (e.g. Beta cases due to small number; non-Gamma cases bearing E484K substitution; or otherwise indeterminate lineages), 2657/2956 (90%) case viruses contributed to variant-specific VE, including: 1332 (50%) Alpha, 1017 (38%) Gamma, 101 (4%) Delta and 207 (8%) non-VOC (of which 76 (37%) were VOIs). Among non-VOC, the B.1.438.1 lineage was most frequent (92/207; 44%) with the Epsilon (B.1.429) VOI contributing next most (70/207; 34%). The Alpha, Gamma, and Delta VOC contribution was similar to the week 17-20 provincial profile overall for adults 50-69 years of age (48%, 34%, and 4%, respectively)[8]. The percentage distribution by VOC status did not vary by >10% (absolute) between participant sub-groups, with the exception of greater Gamma contribution within VCHA versus FHA or other HAs (53%, 32%, and 18%) (**Supplementary Figure 1**). Information on international travel was available for 2618/2657 (99%) cases included in variant-specific VE analyses but just 14 (0.5%) reported such travel (11 Alpha, 2 Gamma, 1 Delta).

### VE estimates

As exemplified in **Supplementary Table 3**, adjusted VE did not vary meaningfully by approach used for control selection. We present VE using all test-negative specimens as controls in **Figures 1-4**, with details provided in **Supplementary Tables 3-8**. The same controls were used for VE against infection and hospitalization.

**Figure 1.**
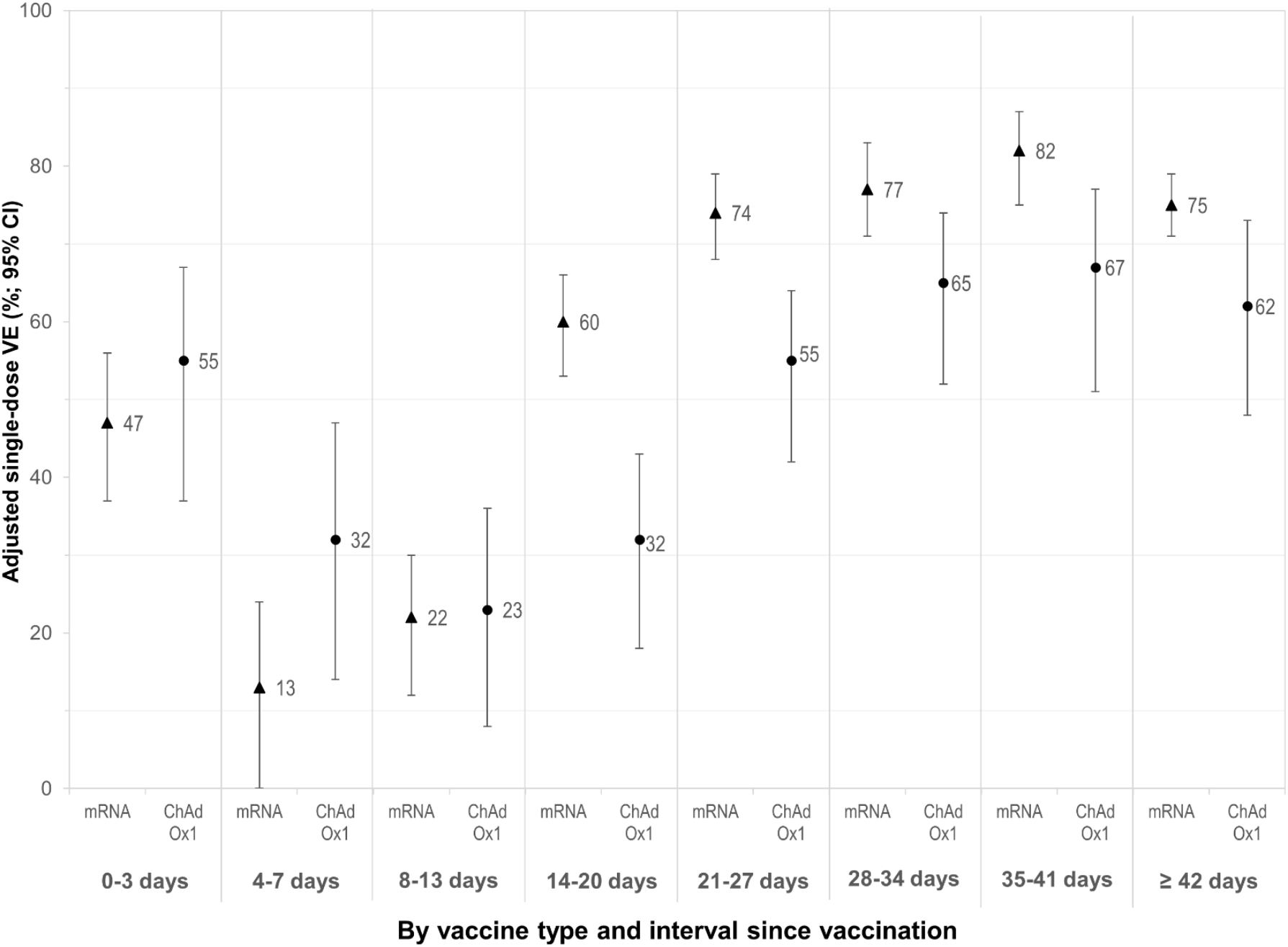
Adjusted vaccine effectiveness against infection by vaccine type and interval in days since vaccination, adults 50-69-years-old, British Columbia, Canada, weeks 14-20 VE = vaccine effectiveness; CI = confidence interval; mRNA = messenger RNA; ChAdOx1 = Chimpanzee adenoviral vectored vaccine All vaccine effectiveness estimates are adjusted for age group (50-59, 60-69 years); gender (men, women); individual epidemiological week (14-20); and health authority (HA) (Fraser HA, Interior HA, Northern HA, Vancouver Coastal HA, Vancouver Island HA). See **Supplementary Table 4** for details.

**Figure 2.**
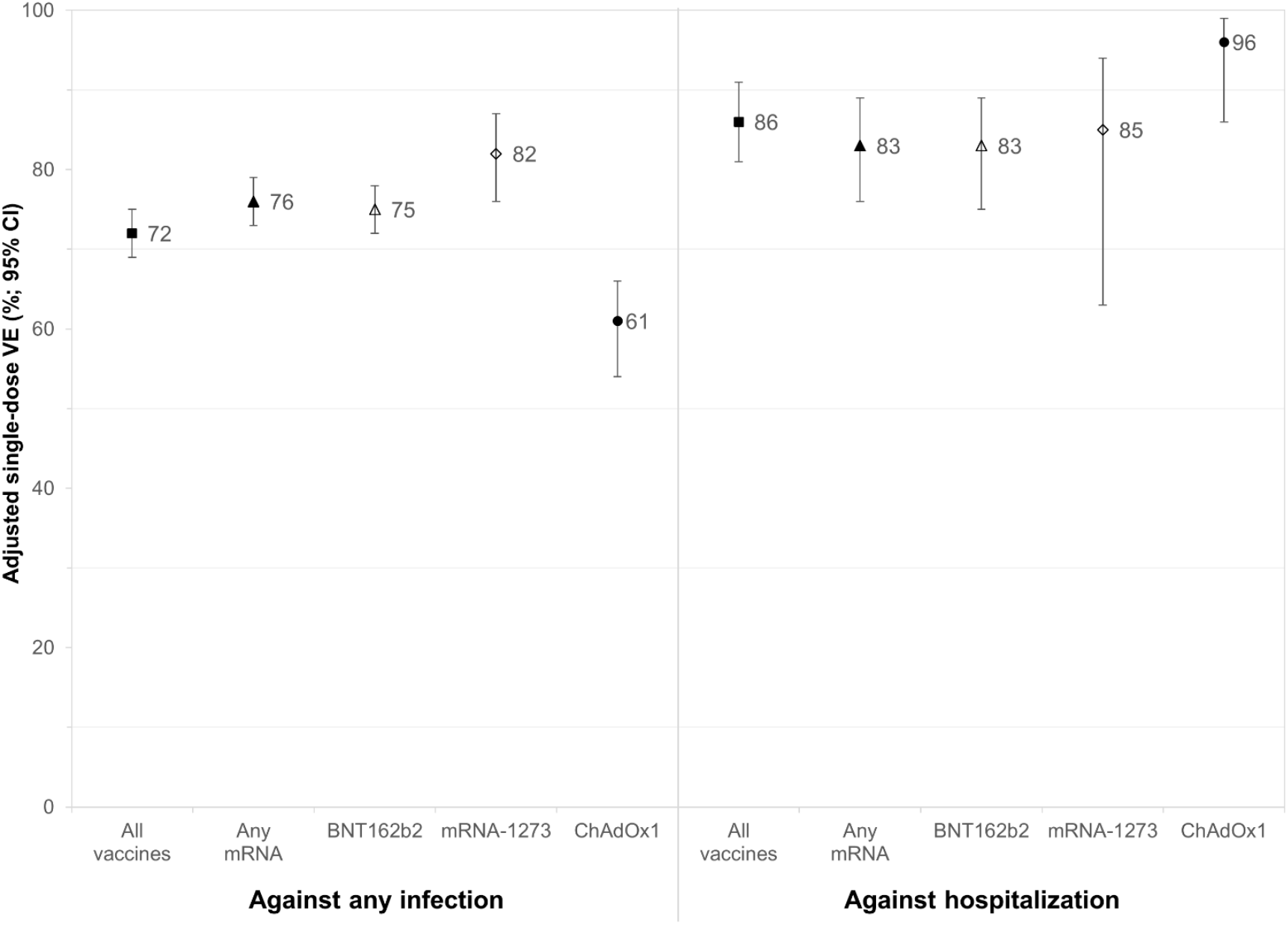
Adjusted vaccine effectiveness against infection or hospitalization, ≥21 days post-vaccination, adults 50-69-years-old, British Columbia, Canada, weeks 14-20 VE = vaccine effectiveness; CI = confidence interval; mRNA = messenger RNA; BNT162b2 = mRNA vaccine (Pfizer-BioNTech); mRNA-1273 = mRNA vaccine (Moderna); ChAdOx1 = Chimpanzee adenoviral vectored vaccine (AstraZeneca/COVISHIELD) All vaccine effectiveness estimates are adjusted for age group (50-59, 60-69 years); gender (men, women); individual epidemiological week (14-20); and health authority (HA) (Fraser HA, Interior HA, Northern HA, Vancouver Coastal HA, Vancouver Island HA). See **Supplementary Tables 4, 5 and 7** for details.

**Figure 3.**
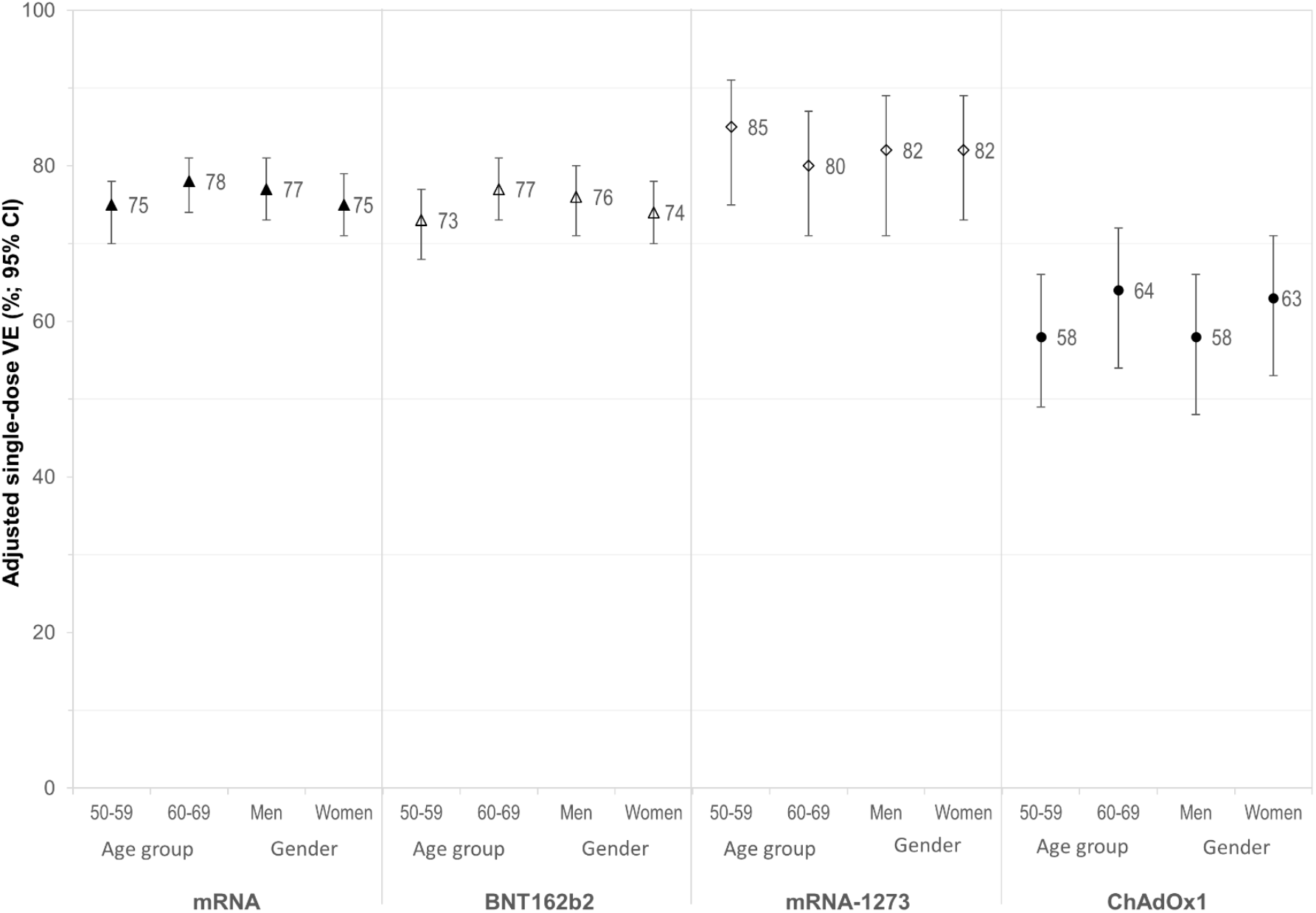
Adjusted vaccine effectiveness against infection ≥21 days post-vaccination by age sub-group and gender, adults 50-69-years-old, British Columbia, Canada, weeks 14-20 VE = vaccine effectiveness; CI = confidence interval; mRNA = messenger RNA; BNT162b2 = mRNA vaccine (Pfizer-BioNTech); mRNA-1273 = mRNA vaccine (Moderna); ChAdOx1 = Chimpanzee adenoviral vectored vaccine (AstraZeneca/COVISHIELD) All vaccine effectiveness estimates are adjusted for individual epidemiological week (14-20); and health authority (HA) (Fraser HA, Interior HA, Northern HA, Vancouver Coastal HA, Vancouver Island HA) and additionally for age group (50-59, 60-69 years) and/or gender (men, women) as indicated. See **Supplementary Table 6** for details.

**Figure 4.**
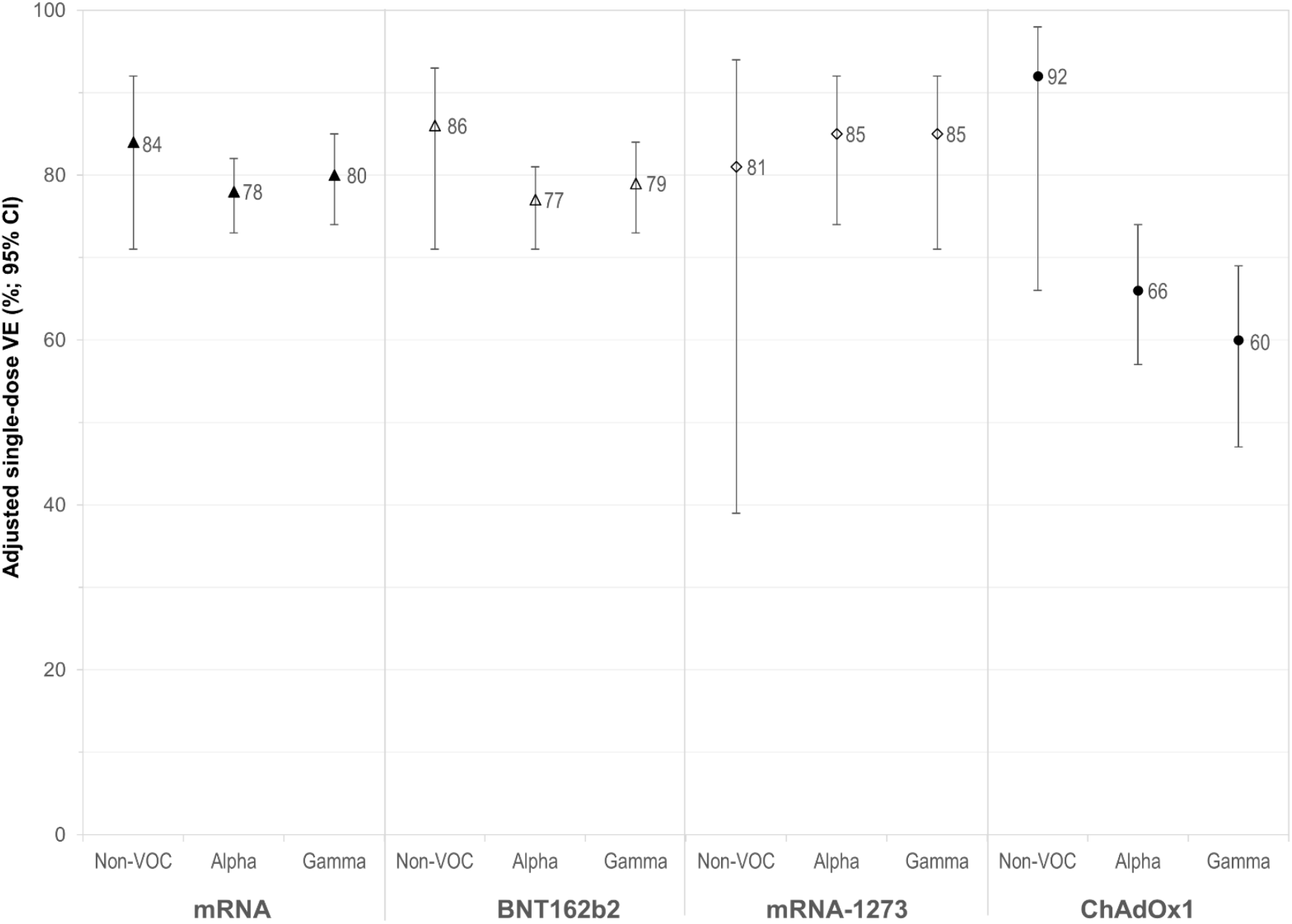
Adjusted vaccine effectiveness against infection due to variants of concern, ≥21 days post-vaccination, adults 50-69-years-old, British Columbia, Canada, weeks 14-20 VE = vaccine effectiveness; CI = confidence interval; mRNA = messenger RNA; BNT162b2 = mRNA vaccine (Pfizer-BioNTech); mRNA-1273 = mRNA vaccine (Moderna); ChAdOx1 = Chimpanzee adenoviral vectored vaccine (AstraZeneca/COVISHIELD); VOC = variant of concern All vaccine effectiveness estimates are adjusted for age group (50-59, 60-69 years); gender (men, women); individual epidemiological week (14-20); and health authority (HA) (Fraser HA, Interior HA, Northern HA, Vancouver Coastal HA, Vancouver Island HA). Owing to sparse data, estimates are not displayed for Delta but for completeness are provided alongside other VOC-specific VE details in **Supplementary Table 8**.

#### VE, by interval since vaccination

At 0-3 days post-vaccination, when there should be no vaccine protection, VE estimates were positively-biased for both mRNA (47%; 95%CI 37-56) and ChAdOx1 (55%; 95%CI 37-67) vaccines (**Figure 1; Supplementary Tables 4-5**). This skew resolved by 4-7 days post-vaccination for mRNA (13%; 95%CI 0-24) but not ChAdOx1 (32%; 95%CI 14-47). By 8-13 days, VE was comparable for mRNA (22%; 95%CI 12-30) and ChAdOx1 (23%; 95%CI 8-36). From 14-20 days, VE increased for both types of vaccine but more steeply for mRNA (60%; 95%CI 53-66) than ChAdOx1 (32%; 95%CI 18-43) and was consistently higher for mRNA vaccines through each weekly interval assessed thereafter.

#### Primary VE, infection and hospitalization

During weeks 14-20, VE against any infection ≥21 days post-vaccination was 76% (95%CI 73-79) for mRNA vaccines: 75% (95%CI 72-78) for BNT162b2 and 82% (95%CI 76-87) for mRNA-1273 (**Figure 2; Supplementary Tables 4-5**). VE for ChAdOx1 was significantly lower than either mRNA vaccine at 61% (95%CI 54-66). VE did not vary by age sub-group or gender for any vaccine (**Figure 3; Supplementary Table 6**) and was similar in VCHA and FHA for both mRNA (74%; 95%CI 67-79 vs. 79%; 95%CI 75-82) and ChAdOx1 (61%; 95%CI 43-73 vs. 64%; 95%CI 57-71).

VE against hospitalization was comparable but slightly higher than against infection for BNT162b2 (83%; 95%CI 76-89) and mRNA-1273 (85%; 95%CI 75-89). VE against hospitalization was significantly higher than against infection for ChAdOx1 (96%; 95%CI 86-99) (**Figure 2; Supplementary Table 7**).

#### VE, by VOC

Restricted to weeks 17-20, VE ≥21 days post-vaccination was similar to the full analysis period for all vaccines and against both infection and hospitalization (**Supplementary Tables 7-8**).

VE was generally highest against non-VOC infections for mRNA (84%; 95%CI 71-92) and ChAdOx1 (92%; 95%CI 66-98) (**Figure 4; Supplementary Table 8**), and was similar for mRNA vaccines against non-VOCs excluding VOIs (87%; 95%CI 71-95) [insufficient sample size for ChAdOx1]. mRNA estimates were driven foremost by BNT162b2 for which VE was 86% (95%CI 71-93) against non-VOC, 77% (95%CI 71-81) against Alpha and 79% (95%CI 73-84) against Gamma. Although lower overall, ChAdOx1 VE was also comparable against Alpha and Gamma infections (66%; 95%CI 57-74 and 60%; 95%CI 48-69). Variant-specific VE against hospitalization was generally 80% or more, recognizing small sample size in this further stratified analysis (**Supplementary Table 7**).

With sparse data, VE estimates against Delta are more unstable and require cautious interpretation. Whereas unadjusted and adjusted VE estimates varied by no more than 5% (absolute) in all other primary analyses, including variant-specific, they differed more against Delta infections for both mRNA (62%; 95%CI 28-80 and 74%;95%CI 47-87) and ChAdOx1 (47%; 95%CI -24-77 and 73%; 95%CI 35-88) vaccines (**Supplementary Table 8**).

## DISCUSSION

In this post-marketing observational study, we directly compared the effectiveness of two major new vaccine innovations (mRNA and vector-based), deployed for the first time on a mass population scale during the COVID-19 pandemic. Among adults 50-69-years-old eligible for both vaccines in BC, Canada, a single dose reduced the overall SARS-CoV-2 infection risk by 76% for mRNA (75% for BNT162b2 and 82% for mRNA-1273), and 61% for ChAdOx1 recipients, with VE against hospitalization exceeding 80% for all vaccines. Such protection is particularly meaningful because it was measured in the midst of a severe spring wave in BC when a variety of newly-emergent VOC were circulating.

Although no head-to-head RCT comparisons of relative efficacy are yet available, the pattern of higher mRNA vs. ChAdOx1 effectiveness we report is consistent with indirect comparison across separate pre-marketing, product-specific RCTs, similarly showing higher protection with a single dose of mRNA (92-93%) vs. ChAdOx1 (76%) vaccine against clinical illness[2-4]. Our point estimate of ChAdOx1 VE against any infection (61%) is moreover very similar to the pooled RCT estimate of ChAdOx1 efficacy against any infection (64%)[4]. The more rapid increase in protection we observed among mRNA recipients also aligns with the faster antibody responses reported for mRNA vs. ChAdOx1 in head-to-head immunogenicity comparison [17]. We qualify the lower estimates of ChAdOx1 efficacy by showing one dose may have been less protective against infection per se, but was at least as protective as mRNA vaccines against severe outcomes. With few hospitalizations among vaccinated participants overall, our product-specific VE estimates against hospitalization were accompanied by wider confidence intervals. However, better ChAdOx1 protection against more severe outcomes also aligns with findings from immunogenicity comparisons, showing lower antibody but comparable or even higher T cell responses with a single dose of ChAdOx1 vs. mRNA vaccine [17,18].

Our study also adds to the understanding of VE against emergent VOC. As recommended to reduce selection bias [19], we confirmed the VOC status of nearly all (∼90%) of the case viruses contributing to variant-specific VE through systematic genome sequencing. Both mRNA and ChAdOx1 vaccines protected best against viruses lacking key spike mutations (such as E484K) that are the hallmarks of some VOC. In that regard, our VE exceeding 80% against non-VOC infections is most comparable to RCT estimates prior to VOC emergence [2-4]. We acknowledge, however, that non-VOCs have become increasingly displaced, reducing their available sample size, and resulting in confidence intervals overlapping VOC estimates. As in our previous analysis of mRNA vaccines among adults ≥70-years-old [11], VE was comparable against Alpha and Gamma variants, here shown for both mRNA and ChAdOx1. In combination, our findings suggest minimal impact of the earliest-emergent Alpha and Gamma VOCs on single-dose vaccine protection. Although we did not identify a particular signal of concern pertaining to Delta, that VOC had not yet established a stronghold in BC during the spring 2021 wave [8,9]. Follow-up analyses are underway, inclusive of younger adults among whom the Delta variant has since more prominently propagated, and across an extended period to enable two-dose comparisons.

In addition to comparison with gold-standard RCT evidence, our findings may also be compared with other observational studies. Elsewhere within Canada, Chung et al. used the TND to estimate VE for the province of Ontario between mid-December, 2020 and mid-April, 2021[20]. In subset analysis of adults 40-69 years, they reported single-dose VE (≥14 days post-vaccination) against symptomatic illness that was lower than we report for BNT162b2 (64% vs. our 75%) but comparable to our finding for mRNA-1273 (both 82%), with VE against hospitalization similarly exceeding 80% for both products. However, their analyses did not include ChAdOx1. In a more recent pre-print from Ontario, Nasreen et al. update to May 2021 to provide variant-specific estimates, including for ChAdOx1[21]. However, their assignment of VOC status was largely based on probabilistic model imputation rather than empirical confirmation through virus gene sequencing, risking misclassification of non-VOC and Delta cases in particular. They also combined analysis of Beta/Gamma variants although these variants may not be comparable in their potential for vaccine escape: reductions in neutralizing antibody for Beta have been more severe and in clinical trials, ChAdOx1 showed no efficacy against mild-moderate Beta illness [22-24]. Among adults <60-years-old, Nasreen et al. reported lower single-dose BNT162b2 VE against non-VOC (65%), Alpha (71%), Beta/Gamma (65%) and Delta (62%) infections. Unlike us, they found no real difference between ChAdOx1 and BNT162b2 against Alpha (67%) or Delta (67%), with the ChAdOx1 VE lower only against Beta/Gamma (43%). The variant-specific VE estimates from both BC and Ontario, Canada are nevertheless aligned in each being higher than reported in TND analysis from the United Kingdom (UK)[25,26]. Among individuals ≥16-years-old in England between October 2020 and May 2021 [25], Bernal et al report comparable BNT162b2 and ChAdOx1 VE against Alpha (48% and 49%, respectively) and Delta (36% and 30%, respectively) that are lower than from Canada and also lower for both vaccines against Alpha than the same authors had reported previously (60-70%)[27,28]. Among individuals ≥15-years-old in Scotland between April and June 2021, Sheikh et al. report similarly low VE for BNT162b2 and ChAdOx1 against Alpha (38% and 37%) and Delta (30% and 18%)[26]. In both UK studies, distinction between Alpha and Delta variants was primarily based upon presumptive S-gene screen, and in comparing across these several studies other differences in target populations and methods should also be considered.

The main limitation of our study, as elsewhere, is reliance on general laboratory submissions and clinical or surveillance data subject to missing information, misclassification, and selection bias. We attempted to standardize the likelihood of test-positivity by excluding specimens collected for non-clinical screening purposes (e.g. private testing). Testing for vaccine-associated adverse events mimicking but not due to COVID-19 (e.g., flu-like symptoms) may explain positively-biased VE estimates during the first few days following vaccination, which persisted longer for ChAdOx1 perhaps owing to heightened vaccine safety concerns. Aligned with that hypothesis, we highlight lower test-positivity among vaccinated individuals during days 0-3 vs. 4-7 post-vaccination (**Supplementary Table 4**). The TND partially standardizes for healthcare seeking behaviours, while other variations associated with both vaccination and exposure risk could still play a role. We cannot rule out residual bias and confounding. Unlike mRNA vaccines, ChAdOx1 was available to eligible age groups in BC through pharmacies without awaiting invitation to public clinics; those at higher risk may have preferentially received this vaccine, contributing to lower VE. Conversely, if ChAdOx1 recipients were generally more risk averse in their behaviours our ChAdOx1 VE may also be an over-estimate. Reassuring against bias in either direction, ChAdOx1 estimates reported here align well with clinical trial findings.

In conclusion, a single dose of mRNA vaccine reduced the SARS-CoV-2 infection risk among adults 50-69-years-old by at least 75%, including due to Alpha and Gamma VOC. Although less protective against any infection, a single dose of ChAdOx1 also reduced the SARS-CoV-2 risk overall by about 60% and protected against severe outcomes at least as well as mRNA vaccines, reducing the hospitalization risk by more than 80%. Robust estimates of single-dose VE against the Delta variant are needed to further inform optimal use of available doses while global vaccine supplies remain constrained.

## Data Availability

Data sharing that complies with relevant legislation to protect privacy and confidentiality will be considered upon request.

## Acknowledgments

Authors thank the following individuals from the BC Centre for Disease Control: Yayuk Joffres for quality control of laboratory data; Yin Chang for laboratory data management; and Samantha Kaweski for laboratory coordination support. We thank the Provincial Public Health Information Systems (PPHIS) team for contributions related to the provincial immunization registry (PIR). Manish Sadarangani acknowledges general salary support provided to him by awards from the BC Children’s Hospital Foundation, the Canadian Child Health Clinician Scientist Program and the Michael Smith Foundation for Health Research. Finally, we thank the many frontline, regional and provincial practitioners, including clinical, laboratory and public health providers, epidemiologists, Medical Health Officers, laboratory staff, vaccinators, participants and others who contributed to the epidemiological, virological and genetic characterization data underpinning these analyses.

## Funding

No external funding was provided for this work.

## Potential conflicts of interest

DMS is Principal or co-Investigator on grants from the Michael Smith Foundation for Health Research, the Public Health Agency of Canada, and the Canadian Institutes of Health Research paid to her institution and unrelated to the current work. MK received grants/contracts paid to his institution from Roche, Hologic and Siemens, unrelated to this work. MS has been an investigator on projects, unrelated to the current work, funded by GlaxoSmithKline, Merck, Pfizer, Sanofi-Pasteur, Seqirus, Symvivo and VBI Vaccines. All funds have been paid to his institute, and he has not received any personal payments. Other authors have no conflicts of interest to disclose.

## Notes

### Clinical Trial

Not applicable

### Author Declarations

Data linkages and analyses were authorized by the Provincial Health Officer under the Public Health Act and exempt from research ethics board review by the University of British Columbia Research Ethics Board.

