## Supplementary_Material for "Comparative single-dose mRNA and ChAdOx1 vaccine effectiveness against SARS-CoV-2, including early variants of concern: a test-negative design, British Columbia, Canada"

**Supplementary Material 1** Genetic characterization of SARS-CoV-2 viruses

To understand the mix of SARS-CoV-2 viruses underpinning vaccine effectiveness (VE) findings and to support variant-specific VE estimates, we attempted to genetically characterize all viruses collected from the 2956 cases 50-69 years old who contributed to VE analyses between epi-weeks 17 and 20 of the study period.

Viruses were genetically grouped based upon: (a) whole genome sequencing (WGS) and/or (b) real-time reverse transcription polymerase chain reaction (RT-PCR) screening assays targeting signature mutations of particular variants of concern (VOC)[a,b,c]. Where WGS and screening RT-PCR assays were discordant, WGS results were generally accepted.

Whole genome sequencing

WGS was performed at the British Columbia Centre for Disease Control Public Health Laboratory (BCCDC PHL). In brief, SARS-CoV-2 RNA was extracted using the Applied BioSystems MagMax™ Express 96 Nucleic Acid Extractor and the MagMax Viral/Pathogen Nucleic Acid Isolation Kit (Thermo Fisher). Viral RNA was reverse transcribed into cDNA using Thermo SuperScript IV. A 1200bp tiled amplicon scheme[d,e] was used to amplify the entire SARS-CoV-2 genome[f]. DNA libraries were prepared using DNA Prep Library Preparation Kit (Illumina) and sequenced on a MiSeq or NextSeq 2000 instrument (Illumina, San Diego). SARS-CoV-2 whole genome consensus sequences were generated using a modified Nextflow pipeline for running the ARTIC network's[g] field bioinformatics tools[h]. Lineages were assigned to the respective VOC/VOI catagories and their decedents using Pangolin (from version 2.4.2, pangoLEARN 2021-05-19 and up)[i].Sequencing quality control metrics and mutational profiles were monitored using nCoV-tools from the Simpson Lab[j].

Screening RT-PCR

Four VOC screening RT-PCR assays were used at the BCCDC PHL and various hospital-based laboratories across the province including: (1) a laboratory-developed N501Y single nucleotide polymorphism (SNP) PCR at the Victoria General Hospital in Vancouver Island Health Authority[k]; (2) a laboratory-developed N501Y and E484K dual SNP PCR at the BCCDC PHL[k]; (3) a laboratory developed sequential deletion and SNP PCR including N501Y and K417T at the St. Paul's Hospital

[a] Government of Canada. SARS-CoV-2 variants: National definitions, classifications and public health actions. [Accessed 29 August 2021]. Available: https://www.canada.ca/en/public-health/services/diseases/2019-novel-coronavirus-infection/health-professionals/testing-diagnosing-case-reporting/sars-cov-2-variants-national-definitions-classifications-public-health-actions.html

[b] World Health Organization. Tracking SARS-CoV-2 variants. [Accessed 20 September 2021]. Available: https://www.who.int/en/activities/tracking-SARS-CoV-2-variants/

[c] Center for Viral Systems Biology (CViSB), Scripps Research. SARS-CoV-2 (hCoV-19) mutation reports. Lineage | Mutation tracker. [Accessed 29 August 2021]. Available: https://outbreak.info/situation-reports

[d] Freed NE, Vlková M, Faisal MB, Silander OK. Rapid and inexpensive whole-genome sequencing of SARS-CoV-2 using 1200 bp tiled amplicons and Oxford Nanopore Rapid Barcoding. Biol Methods Protoc. 2020 Jul 18;5(1):bpaa014.

[e] Kuchinski KS, Nguyen J, Lee TD, et al. Mutations in emerging variant of concern lineages disrupt genomic sequencing of SARS-CoV-2 clinical specimens. MedRxiv 2021.06.01.21258181; (Pre-print) doi: https://doi.org/10.1101/2021.06.01.21258181

[f] Galloway SE, Paul P, MacCannell DR, et al. Emergence of SARS-CoV-2 B.1.1.7 Lineage - United States, December 29, 2020-January 12, 2021. MMWR Morb Mortal Wkly Rep. 2021 Jan 22;70(3):95-9.

[g] https://artic.network/ncov-2019

[h] https://github.com/BCCDC-PHL/ncov2019-artic-nf

[i] Rambaut A, Holmes EC, O'Toole A, et al. A dynamic nomenclature proposal for SARS-CoV-2 lineages to assist genomic epidemiology. Nature Microbiology 2020;5:1403-07. Available: https://www.nature.com/articles/s41564-020-0770-5

[j] https://github.com/jts/ncov-tools

[k] Hogan CA, Jassem AN, Sbihi H, et al. Rapid increase in SARS-CoV_2 P.1 lineage leading to codominance with B.1.1.7 lineage, British Columbia, Canada, January-April 2021. Emerging Infectious Diseases 2021;27: [early release]. Doi: 10.3201/eid2711.211190. Available: https://wwwnc.cdc.gov/eid/article/27/11/21-1190_article

Laboratory in Vancouver Coastal Health Authority[a]; and (4) the commercial Seegene Allplex™ SARS-CoV-2 Variant I Assay (Seegene, Seoul, South Korea) targeting N501Y and E484K at the Kelowna General Hospital Microbiology Laboratory in Interior Health Authority. Based on these methods, viruses were genetically characterized/categorized as per the algorithm below:

**Alpha (Pango lineage: B.1.1.7)**
WGS = B.1.1.7 or
Screening RT-PCR = Positive for N501Y and positive for 69/70 deletion or positive for N501Y and negative for E484K (presumptive)

**Beta (Pango lineage: B.1.351)**
WGS = B.1.351 or
Screening RT-PCR = Positive for N501Y and negative for K417T (presumptive)

**Delta (Pango lineage: B.1.617.2)**
WGS = B.1.617.2
Screening RT-PCR = Not established

**Gamma (Pango lineage: P.1)**
WGS = P.1 or
Screening RT-PCR = Positive for N501Y and positive for K417T (presumptive)

Cases with unavailable or insufficient specimen for genetic characterization were excluded. Taking into account the potentially pivotal 484K substitution and circumstances where limited mutational information could not confirm a lineage, we also excluded from variant-specific analyses:

a. All viruses with the 484K substitution that were identified by WGS to be: (1) the Beta lineage (owing to limited number for VE estimation); or (2) not the Gamma lineage (because atypical of other variants but potentially influential on VE regardless of lineage);
b. All viruses with the 484K substitution but missing WGS results that were: (1) 501Y positive but missing 417 SNP testing (because not possible to distinguish between Beta and Gamma variants; or (2) 501Y negative (because potentially influential on VE estimates if included as a non-VOC and not otherwise classifiable genetically);
c. All viruses without the 484K substitution and missing WGS results that were: (1) 501Y negative (because absent WGS, unable to rule out the Delta variant or Kappa or other VOI);
d. All viruses missing information on the 484K substitution and missing WGS results that were: (1) 501Y positive (because potentially influential on VE estimates, including among viruses with this profile not otherwise classifiable genetically as a VOC); and (2) 501Y negative (because absent WGS, unable to rule out the Delta variant or Kappa or other VOI).

Taking the above into account, remaining viruses were considered non-VOC and either variants of interest (VOI) or non-VOI[b,c,d], with the analysis of VE for non-VOC including non-VOI in the primary analysis but excluding non-VOI in sensitivity analysis.

The ultimate tally by genetic subgroup and final categorizations for the purpose of variant-specific VE analyses, with rationale provided in footnotes, are shown in **Supplementary Table 1** below.

[a] Matic N, Lowe CF, Ritchie G, et al. Rapid detection of SARS-CoV-2 variants of concern, including B.1.1.28/P.1, British Columbia, Canada. 2021;27(6):1673-1676. doi:10.3201/eid2706.210532.https://wwwnc.cdc.gov/eid/article/27/6/21-0532_article
[b] Government of Canada. SARS-CoV-2 variants: National definitions, classifications and public health actions. [Accessed 29 August 2021]. Available: https://www.canada.ca/en/public-health/services/diseases/2019-novel-coronavirus-infection/health-professionals/testing-diagnosing-case-reporting/sars-cov-2-variants-national-definitions-classifications-public-health-actions.html
[c] World Health Organization. Tracking SARS-CoV-2 variants. [Accessed 20 September 2021]. Available: https://www.who.int/en/activities/tracking-SARS-CoV-2-variants/
[d] Center for Viral Systems Biology (CViSB), Scripps Research. SARS-CoV-2 (hCoV-19) mutation reports. Lineage | Mutation tracker. [Accessed 29 August 2021]. Available: https://outbreak.info/situation-reports

**Supplementary Table 1:** Tally of characterized study viruses, by genetic sub-group

| RT-PCR test-positive cases, VE analyses Adults 50-69 years old, epi-weeks 17-20 | Categorization for Variant-specific VE Analysis | Tally (N=2956) |
|---|---|---|
| **Characterized and included in variant-specific VE analyses** | | **2657 (90%)** |
| **Total VOC** | | **2450** |
| Alpha (B.1.1.7) | Alpha | 1332[a] |
| Gamma (P.1) | Gamma | 1017[b] |
| Delta (B.1.617.2) | Delta | 101[c] |
| **Total Non-VOC/non-VOI** | | **131[c]** |
| B.1.438.1 | Non-VOC | 92 |
| B.1.2 | | 28 |
| B.1.480 | | 5 |
| B.1.577 | | 2 |
| B.1.438 | | 1 |
| B.1.9.5 | | 2 |
| AL.1 | | 1 |
| **Total Non-VOC/VOI** | | **76[c]** |
| Epsilon (B.1.429) | Non-VOC[d] | 70 |
| Kappa (B.1.617.1) | Non-VOC[d] | 6 |
| **Characterized but excluded from variant-specific VE analyses** | | **141 (5%)** |
| **Genetic sub-group determined (but non-Gamma variant, 484K positive)** | | **26** |
| Alpha (B.1.1.7; VOC) | Excluded | 5 |
| Beta (B.1.351; VOC) | | 4[e] |
| Eta (B.1.525; Non-VOC / VOI) | | 11 |
| B.1.1.318 (Non-VOC / VOI) | | 4 |
| B.1.438.1 (Non-VOC / Non-VOI) | | 2 |
| **Genetic sub-group indeterminate (and missing WGS)** | | **115** |
| 484K positive | Excluded | 53 |
| 501Y positive, missing 417 SNP[f] | | 52 |
| 501Y negative[g] | | 1 |
| 484K negative | | 32 |
| 501Y negative[h] | | 32 |
| 484K missing | | 30 |
| 501Y positive[i] | | 17 |
| 501Y negative[h] | | 13 |
| **Not characterized and excluded: sample unavailable or insufficient** | | **158 (5%)** |

VE = vaccine effectiveness; VOC = variant of concern; VOI = variant of interest; WGS = whole genome sequencing; SNP = single nucleotide polymorphism

[a] Includes 1072 characterized by WGS and another 260 presumptively characterized by screening assay.
[b] Includes 903 characterized by WGS and another 114 presumptively characterized by screening assay
[c] All characterized by WGS.
[d] Also excluded from variant-specific VE estimation in sensitivity analyses
[e] Excluded from VOC-specific VE analysis because Beta viruses were too few for that separate analysis (and also 484K positive)
[f] Excluded from variant-specific VE analyses because absent information on the 417 SNP it is not possible to distinguish the Beta and Gamma variants among viruses with this profile.
[g] Excluded from variant-specific VE analyses because the 484K substitution is considered pivotal and may be influential on VE if included as a non-VOC (and otherwise not classifiable genetically)
[h] Excluded from variant-specific VE analyses because absent WGS, unable to rule out Delta variant or Kappa or other VOI
[i] Excluded from variant-specific VE analyses because the 484K substitution is considered pivotal and may be influential on VE and not otherwise classifiable genetically as a VOC

**Supplementary Figure 1:** Distribution of variants of concern (VOC) among study viruses, adults 50-69 years old, British Columbia (BC), Canada, epidemiological weeks 17-20

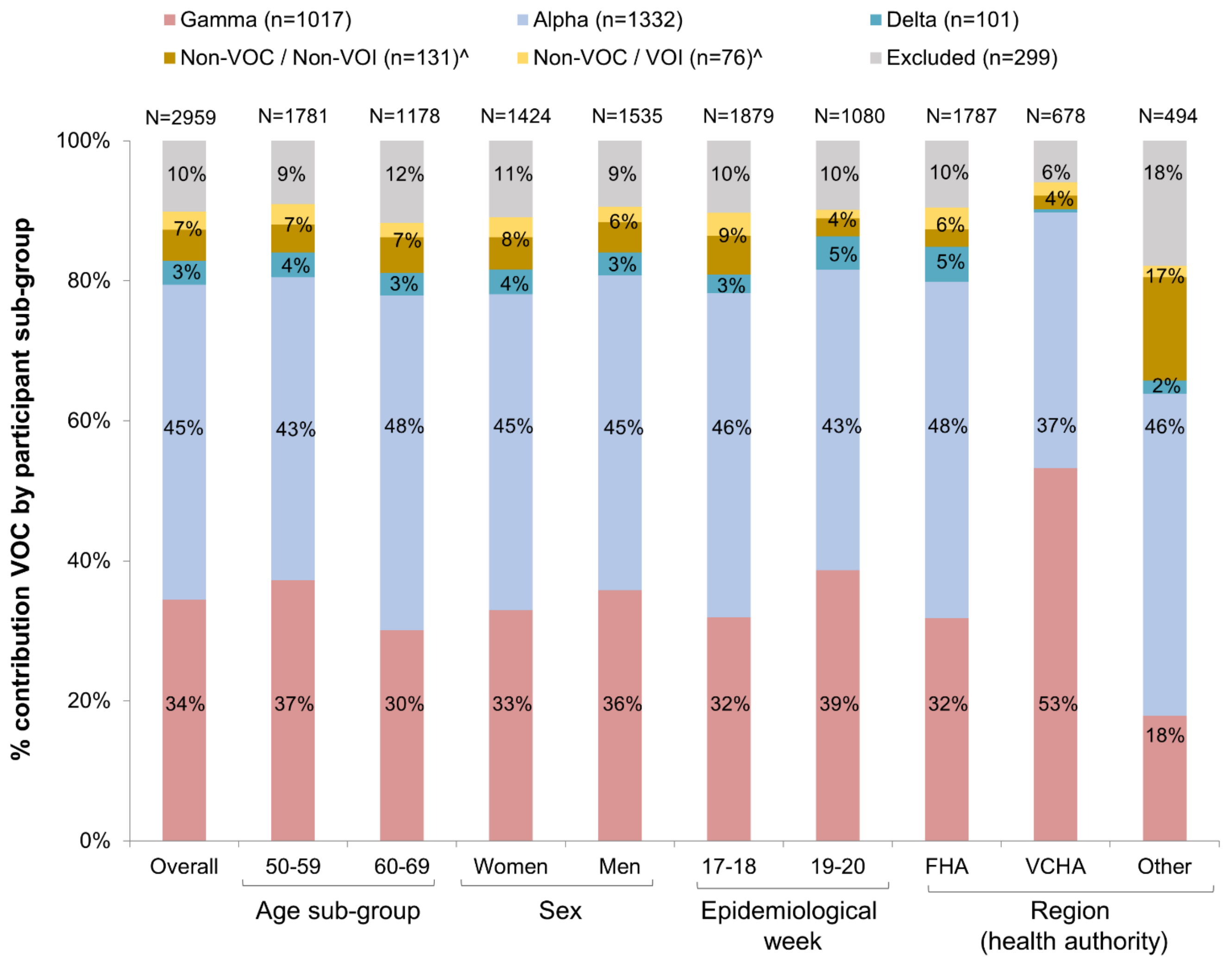

VOC = variant of concern; VOI = variant of interest; for details on genetic characterization and sub-grouping see **Supplementary Table 1**.

^ In primary variant-specific vaccine effectiveness (VE) analyses, non-VOC analysis combines non-VOI and VOI viruses together; accordingly, their combined percentages are displayed here. In sensitivity VE analysis, the non-VOC category excluded VOIs.

**Supplementary Figure 2**. Flowchart of specimens from adults 50-69 years old included in vaccine effectiveness (VE) analyses, April 4 (week 14) to May 22 (week 20), 2021, British Columbia (BC), Canada

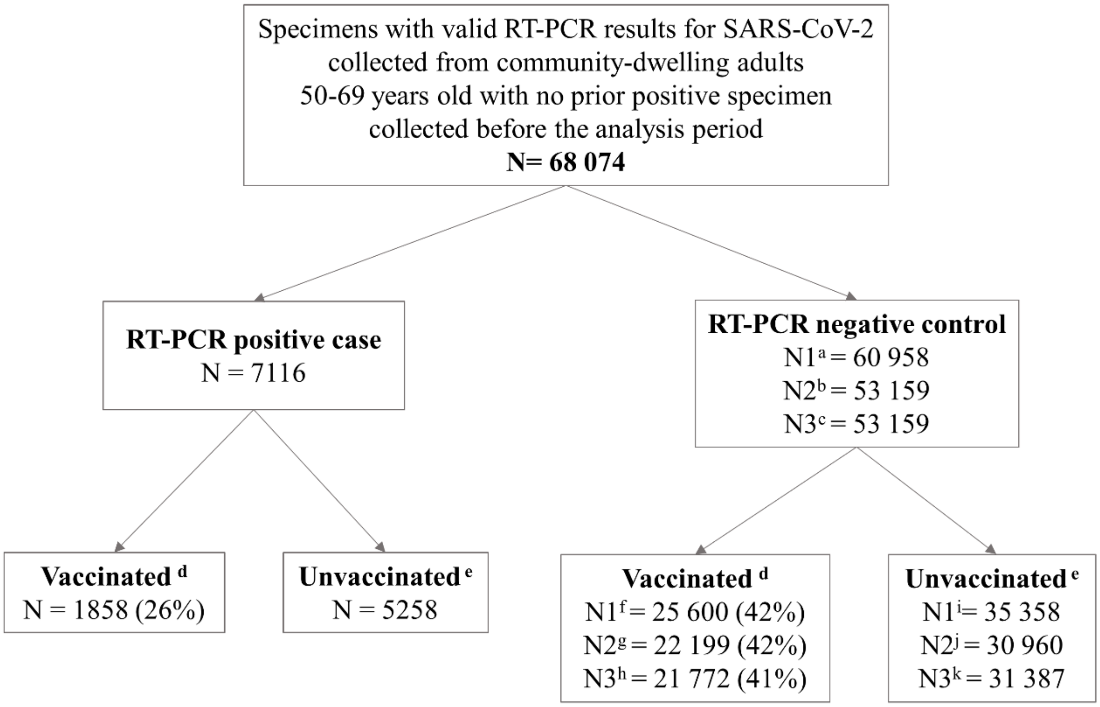

a. Approach 1 for control selection: all negative specimens collected from individuals before the end of the analysis period or becoming a case, whichever occurs first
b. Approach 2 for control selection: one negative specimen (the latest) collected per individual before the end of the analysis period or becoming a case, whichever occurs first
c. Approach 3 for control selection: one randomly selected specimen collected per individual before the end of the analysis period or becoming a case, whichever occurs first
d. Vaccinated on or before the date of specimen collection
e. Not vaccinated on or before the date of specimen collection
f. Vaccinated controls selected as per approach 1
g. Vaccinated controls selected as per approach 2
h. Vaccinated controls selected as per approach 3
i. Unvaccinated controls selected as per approach 1
j. Unvaccinated controls selected as per approach 2
k. Unvaccinated controls selected as per approach 3

**Supplementary Table 2.** Characteristics of vaccinated participants by vaccine type, adults 50-69 years, BC, Canada

| Characteristic | Vaccinated[a] participants overall | | Vaccinated[a] test-positive cases | | Vaccinated[a] test-negative controls[b] | |
|---|---|---|---|---|---|---|
| | mRNA[c], N (%) | ChAdOx1, N (%) | mRNA[c], N (%) | ChAdOx1, N (%) | mRNA[c], N (%) | ChAdOx1, N (%) |
| Number of vaccinated participants | 20 479 | 6979 | 1282 | 576 | 19 197 | 6403 |
| **Age group (years)** | | | | | | |
| 50-59 | 9240 (45) | 4058 (58) | 602 (47) | 317 (55) | 8638 (45) | 3741 (58) |
| 60-69 | 11 239 (55) | 2921 (42) | 680 (53) | 259 (45) | 10 559 (55) | 2662 (42) |
| Median age | 61 | 58 | 60 | 59 | 61 | 58 |
| **Gender** | | | | | | |
| Male | 8980 (44) | 3559 (51) | 632 (49) | 329 (57) | 8348 (43) | 3230 (50) |
| Female | 11 499 (56) | 3420 (49) | 650 (51) | 247 (43) | 10 849 (57) | 3173 (50) |
| **Epidemiological week of specimen collection** | | | | | | |
| 14 | 1489 (7) | 440 (6) | 89 (7) | 55 (10) | 1400 (7) | 385 (6) |
| 15 | 1945 (9) | 779 (11) | 132 (10) | 98 (17) | 1813 (9) | 681 (11) |
| 16 | 2419 (12) | 989 (14) | 173 (13) | 85 (15) | 2246 (12) | 904 (14) |
| 17 | 3208 (16) | 1301 (19) | 208 (16) | 97 (17) | 3000 (16) | 1204 (19) |
| 18 | 3779 (18) | 1391 (20) | 239 (19) | 123 (21) | 3540 (18) | 1268 (20) |
| 19 | 3851 (19) | 1171 (17) | 266 (21) | 76 (13) | 3585 (19) | 1095 (17) |
| 20 | 3788 (18) | 908 (13) | 175 (14) | 42 (7) | 3613 (19) | 866 (14) |
| **Health authority (HA)** | | | | | | |
| Fraser (FHA) | 10 963 (54) | 4757 (68) | 760 (59) | 412 (72) | 10 203 (53) | 4345 (68) |
| Interior (IHA) | 2871 (14) | 324 (5) | 120 (9) | 21 (4) | 2751 (14) | 303 (5) |
| Northern (NHA) | 674 (3) | 87 (1) | 56 (4) | 8 (1) | 618 (3) | 79 (1) |
| Vancouver Coastal (VCHA) | 4239 (21) | 1462 (21) | 313 (24) | 125 (22) | 3926 (20) | 1337 (21) |
| Vancouver Island (VIHA) | 1732 (8) | 349 (5) | 33 (3) | 10 (2) | 1699 (9) | 339 (5) |
| **Days since vaccination (DSV)** | | | | | | |
| 0-13 | 7398 (36) | 2505 (36) | 762 (59) | 257 (45) | 6636 (35) | 2248 (35) |
| 14-20 | 3148 (15) | 1292 (19) | 179 (14) | 131 (23) | 2969 (15) | 1161 (18) |
| 21-27 | 2482 (12) | 1091 (16) | 94 (7) | 75 (13) | 2388 (12) | 1016 (16) |
| 28-34 | 1968 (10) | 851 (12) | 64 (5) | 46 (8) | 1904 (10) | 805 (13) |
| 35-41 | 1520 (7) | 559 (8) | 41 (3) | 28 (5) | 1479 (8) | 531 (8) |
| 42-55 | 2043 (10) | 598 (9) | 65 (5) | 31 (5) | 1978 (10) | 567 (9) |
| 56-69 | 1087 (5) | 83 (1) | 48 (4) | 8 (1) | 1039 (5) | 75 (1) |
| 70+ | 833 (4) | 0 | 29 (2) | 0 | 804 (4) | 0 |
| Median DSV (range) | 20 (0-143) | 19 (0-65) | 11 (0-111) | 15 (0-61) | 20 (0-143) | 19 (0-65) |

[a] Single-dose recipients only without regard to interval between vaccination and specimen collection.
[b] As per Approach 1 for control selection: includes all test-negative specimens.
[c] Profiles similar for BNT162b2 (Pfizer-BioNTech) and mRNA-1273 (Moderna) recipients and are therefore combined into a single mRNA category

**Supplementary Table 3.** Single-dose VE overall by approach for test-negative control selection, adults 50-69 years, BC, Canada

| Interval (DSV) | Case status | By approach for selecting among repeat test-negative control specimens | | | | | | | | | | | |
|---|---|---|---|---|---|---|---|---|---|---|---|---|---|
| | | Approach 1 (all test-negative controls) | | | | Approach 2 (single latest test-negative control) | | | | Approach 3 (single random test-negative control) | | | |
| | | Vacc N (%) | Unvacc N | Crude VE % (95% CI) | **Adjusted [a] VE % (95% CI)** | Vacc N (%) | Unvacc N | Crude VE % (95% CI) | **Adjusted [a] VE % (95% CI)** | Vacc N (%) | Unvacc N | Crude VE % (95% CI) | **Adjusted [a] VE % (95% CI)** |
| 0-3 | Case | 174 (3) | 5258 | 44 | **49** | 174 (3) | 5258 | 45 | **50** | 174 (3) | 5258 | 45 | **50** |
| | Control | 2107 (6) | 35 358 | (35, 53) | **(40, 56)** | 1868 (6) | 30 960 | (36, 53) | **(41, 57)** | 1873 (6) | 31 387 | (36, 53) | **(41, 57)** |
| 4-7 | Case | 343 (6) | 5258 | 13 | **18** | 343 (6) | 5258 | 14 | **19** | 343 (6) | 5258 | 14 | **19** |
| | Control | 2646 (7) | 35 358 | (2, 22) | **(8, 27)** | 2358 (7) | 30 960 | (4, 24) | **(8, 28)** | 2347 (7) | 31 387 | (4, 24) | **(8, 28)** |
| 8-13 | Case | 502 (9) | 5258 | 18 | **22** | 502 (9) | 5258 | 20 | **22** | 502 (9) | 5258 | 20 | **22** |
| | Control | 4131 (10) | 35 358 | (10, 26) | **(14, 30)** | 3672 (11) | 30 960 | (11, 27) | **(14, 30)** | 3665 (10) | 31 387 | (11, 27) | **(14, 30)** |
| 0-13 | Case | 1019 (16) | 5258 | 23 | **28** | 1019 (16) | 5258 | 24 | **29** | 1019 (16) | 5258 | 23 | **29** |
| | Control | 8884 (20) | 35 358 | (18, 28) | **(23, 33)** | 7898 (20) | 30 960 | (18, 29) | **(23, 34)** | 7885 (20) | 31 387 | (17, 28) | **(24, 34)** |
| 14-20 | Case | 310 (6) | 5258 | 50 | **51** | 310 (6) | 5258 | 49 | **49** | 310 (6) | 5258 | 49 | **49** |
| | Control | 4130 (10) | 35 358 | (43, 55) | **(45, 57)** | 3560 (10) | 30 960 | (42, 55) | **(43, 55)** | 3560 (10) | 31 387 | (42, 55) | **(43, 55)** |
| 21-27 | Case | 169 (3) | 5258 | 67 | **68** | 169 (3) | 5258 | 67 | **67** | 169 (3) | 5258 | 67 | **67** |
| | Control | 3404 (9) | 35 358 | (61, 71) | **(62, 73)** | 2989 (9) | 30 960 | (61, 72) | **(61, 72)** | 2909 (8) | 31 387 | (61, 72) | **(61, 72)** |
| 28-34 | Case | 110 (2) | 5258 | 73 | **73** | 110 (2) | 5258 | 72 | **72** | 110 (2) | 5258 | 72 | **72** |
| | Control | 2709 (7) | 35 358 | (67, 77) | **(68, 78)** | 2343 (7) | 30 960 | (66, 77) | **(66, 77)** | 2255 (7) | 31 387 | (66, 77) | **(66, 77)** |
| 35-41 | Case | 69 (1) | 5258 | 77 | **78** | 69 (1) | 5258 | 76 | **76** | 69 (1) | 5258 | 76 | **76** |
| | Control | 2010 (5) | 35 358 | (71, 82) | **(71, 83)** | 1709 (5) | 30 960 | (70, 81) | **(69, 81)** | 1655 (5) | 31 387 | (70, 81) | **(69, 81)** |
| ≥42 | Case | 181 (3) | 5258 | 73 | **73** | 181 (3) | 5258 | 71 | **70** | 181 (3) | 5258 | 71 | **70** |
| | Control | 4463 (11) | 35 358 | (68, 77) | **(69, 77)** | 3700 (11) | 30 960 | (66, 75) | **(65, 75)** | 3508 (10) | 31 387 | (66, 75) | **(65, 75)** |
| ≥21 | Case | 529 (9) | 5258 | 72 | **72** | 529 (9) | 5258 | 71 | **70** | 529 (9) | 5258 | 71 | **70** |
| | Control | 12 586 (26) | 35 358 | (69, 74) | **(69, 75)** | 10 741 (26) | 30 960 | (68, 74) | **(67, 73)** | 10 327 (25) | 31 387 | (68, 74) | **(67, 73)** |

DSV = Days since vaccination, the interval in days between the date of receipt of the first mRNA dose and the date of specimen collection
Vacc = vaccinated; Unvacc = Unvaccinated; VE = vaccine effectiveness; 95% CI = 95% confidence interval

VE interpretation foremost predicated upon an interval of 21 days since vaccination (yellow highlighted row).

[a] VE estimates adjusted for age group (50-59, 60-69 years); gender (men, women); epidemiological week (14, 15, 16, 17, 18, 19, 20); and health authority (HA) (Fraser HA, Interior HA, Northern HA, Vancouver Coastal HA, Vancouver Island HA).

**Supplementary Table 4.** Single-dose vaccine effectiveness by vaccine type, adults 50-69 years, British Columbia, Canada

| Interval (DSV) | Case status | By vaccine type | | | | | | | |
|---|---|---|---|---|---|---|---|---|---|
| | | mRNA products | | | | ChAdOx1 products | | | |
| | | Vaccinated N (%) | Unvaccinated N | Crude VE % (95% CI) | **Adjusted [a] VE % (95% CI)** | Vaccinated N (%) | Unvaccinated N | Crude VE % (95% CI) | **Adjusted [a] VE % (95% CI)** |
| 0-3 | Case | 135 (3) | 5258 | 44 (33, 53) | **47 (37, 56)** | 39 (1) | 5258 | 47 (27, 62) | **55 (37, 67)** |
| | Control | 1610 (4) | 35358 | | | 497 (1) | 35358 | | |
| 4-7 | Case | 267 (5) | 5258 | 10 (-3, 21) | **13 (0, 24)** | 76 (1) | 5258 | 22 (1, 39) | **32 (14, 47)** |
| | Control | 1990 (5) | 35358 | | | 656 (2) | 35358 | | |
| 8-13 | Case | 360 (6) | 5258 | 20 (11, 29) | **22 (12, 30)** | 142 (3) | 5258 | 13 (-4, 27) | **23 (8, 36)** |
| | Control | 3036 (8) | 35358 | | | 1095 (3) | 35358 | | |
| 0-13 | Case | 762 (13) | 5258 | 23 (16, 29) | **22 (12, 30)** | 257 (5) | 5258 | 23 (12, 33) | **33 (23, 41)** |
| | Control | 6636 (16) | 35358 | | | 2248 (6) | 35358 | | |
| 14-20 | Case | 179 (3) | 5258 | 59 (53, 65) | **60 (53, 66)** | 131 (2) | 5258 | 24 (9, 37) | **32 (18, 43)** |
| | Control | 2969 (8) | 35358 | | | 1161 (3) | 35358 | | |
| 21-27 | Case | 94 (2) | 5258 | 74 (67, 79) | **74 (68, 79)** | 75 (1) | 5258 | 50 (37, 61) | **55 (42, 64)** |
| | Control | 2388 (6) | 35358 | | | 1016 (3) | 35358 | | |
| 28-34 | Case | 64 (1) | 5258 | 77 (71, 82) | **77 (71, 83)** | 46 (1) | 5258 | 62 (48, 71) | **65 (52, 74)** |
| | Control | 1904 (5) | 35358 | | | 805 (2) | 35358 | | |
| 35-41 | Case | 41 (1) | 5258 | 81 (75, 86) | **82 (75, 87)** | 28 (1) | 5258 | 65 (48, 76) | **67 (51, 77)** |
| | Control | 1479 (4) | 35358 | | | 531 (1) | 35358 | | |
| ≥42 | Case | 142 (3) | 5258 | 75 (70, 79) | **75 (71, 79)** | 39 (1) | 5258 | 59 (43, 70) | **62 (48, 73)** |
| | Control | 3821 (10) | 35358 | | | 642 (2) | 35358 | | |
| **≥21** | Case | 341 (6) | 5258 | 76 (73, 79) | **76 (73, 79)** | 188 (3) | 5258 | 58 (51, 64) | **61 (54, 66)** |
| | Control | 9592 (21) | 35358 | | | 2994 (8) | 35358 | | |

DSV = Days since vaccination, the interval in days between the date of receipt of the first mRNA dose and the date of specimen collection
VE = vaccine effectiveness; 95% CI = 95% confidence interval
VE interpretation foremost predicated upon an interval of 21 days since vaccination (yellow highlighted row).

[a] VE estimates adjusted for age group (50-59, 60-69 years); gender (men, women); epidemiological week (14, 15, 16, 17, 18, 19, 20); and health authority (HA) (Fraser HA, Interior HA, Northern HA, Vancouver Coastal HA, Vancouver Island HA).

**Supplementary Table 5.** Single-dose vaccine effectiveness by mRNA vaccine type, adults 50-69 years, British Columbia, Canada

| Interval (DSV) | Case status | By mRNA product | | | | | | | | | | | |
|---|---|---|---|---|---|---|---|---|---|---|---|---|---|
| | | mRNA combined products | | | | BNT162b2 (Pfizer-BioNTech) | | | | mRNA-1273 (Moderna) | | | |
| | | Vacc N (%) | Unvacc N | Crude VE % (95% CI) | **Adjusted [a] VE % (95% CI)** | Vacc N (%) | Unvacc N | Crude VE % (95% CI) | **Adjusted [a] VE % (95% CI)** | Vacc N (%) | Unvacc N | Crude VE % (95% CI) | **Adjusted [a] VE % (95% CI)** |
| 0-3 | Case | 135 (3) | 5258 | 44 | **47** | 100 (2) | 5258 | 44 | **47** | 35 (1) | 5258 | 41 | **47** |
| | Control | 1610 (4) | 35358 | (33, 53) | **(37, 56)** | 1208 (3) | 35358 | (32, 55) | **(35, 57)** | 402 (1) | 35358 | (17, 59) | **(25, 62)** |
| 4-7 | Case | 267 (5) | 5258 | 10 | **13** | 200 (4) | 5258 | 14 | **16** | 67 (1) | 5258 | -8 | **0** |
| | Control | 1990 (5) | 35358 | (-3, 21) | **(0, 24)** | 1572 (4) | 35358 | (1, 26) | **(2, 28)** | 418 (1) | 35358 | (-40, 17) | **(-30, 23)** |
| 8-13 | Case | 360 (6) | 5258 | 20 | **22** | 282 (5) | 5258 | 20 | **21** | 78 (1) | 5258 | 20 | **22** |
| | Control | 3036 (8) | 35358 | (11, 29) | **(12, 30)** | 2379 (6) | 35358 | (10, 30) | **(10, 31)** | 657 (2) | 35358 | (-1, 37) | **(1, 39)** |
| 0-13 | Case | 762 (13) | 5258 | 23 | **22** | 582 (10) | 5258 | 24 | **26** | 180 (3) | 5258 | 18 | **23** |
| | Control | 6636 (16) | 35358 | (16, 29) | **(12, 30)** | 5159 (13) | 35358 | (17, 31) | **(19, 33)** | 1477 (4) | 35358 | (4, 30) | **(10, 35)** |
| 14-20 | Case | 179 (3) | 5258 | 59 | **60** | 149 (3) | 5258 | 55 | **56** | 30 (1) | 5258 | 72 | **73** |
| | Control | 2969 (8) | 35358 | (53, 65) | **(53, 66)** | 2248 (6) | 35358 | (47, 62) | **(48, 63)** | 721 (2) | 35358 | (60, 81) | **(61, 81)** |
| 21-27 | Case | 94 (2) | 5258 | 74 | **74** | 82 (2) | 5258 | 71 | **71** | 12 (<1) | 5258 | 84 | **84** |
| | Control | 2388 (6) | 35358 | (67, 79) | **(68, 79)** | 1889 (5) | 35358 | (64, 77) | **(64, 77)** | 499 (1) | 35358 | (71, 91) | **(72, 91)** |
| 28-41[b] | Case | 105 (2) | 5258 | 79 | **79** | 91 (2) | 5258 | 77 | **78** | 14 (<1) | 5258 | 86 | **86** |
| | Control | 3383 (9) | 35358 | (75, 83) | **(75, 83)** | 2704 (7) | 35358 | (72, 82) | **(73, 82)** | 679 (2) | 35358 | (76, 92) | **(76, 92)** |
| ≥42 | Case | 142 (3) | 5258 | 75 | **75** | 126 (2) | 5258 | 75 | **76** | 16 (<1) | 5258 | 76 | **74** |
| | Control | 3821 (10) | 35358 | (70, 79) | **(71, 79)** | 3366 (9) | 35358 | (70, 79) | **(71, 80)** | 455 (1) | 35358 | (61, 86) | **(57, 84)** |
| **≥21** | Case | 341 (6) | 5258 | 76 | **76** | 299 (5) | 5258 | 75 | **75** | 42 (1) | 5258 | 83 | **82** |
| | Control | 9592 (21) | 35358 | (73, 79) | **(73, 79)** | 7959 (18) | 35358 | (72, 78) | **(72, 78)** | 1633 (4) | 35358 | (76, 87) | **(76, 87)** |

DSV = Days since vaccination, the interval in days between the date of receipt of the first mRNA dose and the date of specimen collection
Vacc = vaccinated; Unvacc = Unvaccinated; VE = vaccine effectiveness; 95% CI = 95% confidence interval
VE interpretation foremost predicated upon an interval of 21 days since vaccination (yellow highlighted row).

[a] VE estimates adjusted for age group (50-59, 60-69 years); gender (men, women); epidemiological week (14, 15, 16, 17, 18, 19, 20); and health authority (HA) (Fraser HA, Interior HA, Northern HA, Vancouver Coastal HA, Vancouver Island HA).

[b] Combined periods (28-34 and 35-41) owing to small sample size when stratified more finely for interval and vaccine type simultaneously, notably with the smaller sample size for the Moderna product

**Supplementary Table 6.** Single-dose vaccine effectiveness (≥21 days since vaccination) by age sub-group and gender, British Columbia, Canada

| Stratum | Case status | Sample sizes | | Vaccine effectiveness (VE) (95% CI) | |
|---|---|---|---|---|---|
| | | Vaccinated N (%) | Unvaccinated N | Crude | **Adjusted** |
| **By age sub-group[a]** | | | | | |
| **50-59 years** | | | | | |
| All vaccines | Case | 275 (8) | 3323 | 70 (66, 73) | **70 (66, 74)** |
| | Control | 6070 (22) | 22 109 | | |
| mRNA vaccines | Case | 168 (5) | 3323 | 75 (70, 78) | **75 (70, 78)** |
| | Control | 4427 (17) | 22 109 | | |
| BNT162b2 (Pfizer-BioNTech) | Case | 154 (4) | 3323 | 73 (68, 77) | **73 (68, 77)** |
| | Control | 3767 (15) | 22 109 | | |
| mRNA-1273 (Moderna) | Case | 14 (<1) | 3323 | 86 (76, 92) | **85 (75, 91)** |
| | Control | 660 (3) | 22 109 | | |
| ChAdOx1 vaccines | Case | 107 (3) | 3323 | 57 (47, 64) | **58 (49, 66)** |
| | Control | 1643 (7) | 22 109 | | |
| **60-69 years** | | | | | |
| All vaccines | Case | 254 (12) | 1935 | 73 (69, 77) | **74 (70, 78)** |
| | Control | 6516 (33) | 13 249 | | |
| mRNA vaccines | Case | 173 (8) | 1935 | 77 (73, 80) | **78 (74, 81)** |
| | Control | 5165 (28) | 13 249 | | |
| BNT162b2 (Pfizer-BioNTech) | Case | 145 (7) | 1935 | 76 (72, 80) | **77 (73, 81)** |
| | Control | 4192 (24) | 13 249 | | |
| mRNA-1273 (Moderna) | Case | 28 (1) | 1935 | 80 (71, 87) | **80 (71, 87)** |
| | Control | 973 (7) | 13 249 | | |
| ChAdOx1 vaccines | Case | 81 (4) | 1935 | 59 (48, 67) | **64 (54, 72)** |
| | Control | 1351 (9) | 13 249 | | |
| **By gender[b]** | | | | | |
| **Men** | | | | | |
| All vaccines | Case | 240 (8) | 2689 | 72 (68, 76) | **72 (67, 76)** |
| | Control | 5412 (24) | 16 868 | | |
| mRNA vaccines | Case | 139 (5) | 2689 | 78 (73, 81) | **77 (73, 81)** |
| | Control | 3910 (19) | 16 868 | | |
| BNT162b2 (Pfizer-BioNTech) | Case | 120 (4) | 2689 | 76 (72, 80) | **76 (71, 80)** |
| | Control | 3187 (16) | 16 868 | | |
| mRNA-1273 (Moderna) | Case | 19 (<1) | 2689 | 84 (74, 90) | **82 (71, 89)** |
| | Control | 723 (4) | 16 868 | | |
| ChAdOx1 vaccines | Case | 101 (4) | 2689 | 58 (48, 66) | **58 (48, 66)** |
| | Control | 1502 (8) | 16 868 | | |
| **Women** | | | | | |
| All vaccines | Case | 289 (10) | 2569 | 71 (67, 74) | **72 (69, 76)** |
| | Control | 7174 (28) | 18 490 | | |
| mRNA vaccines | Case | 202 (7) | 2569 | 74 (70, 78) | **75 (71, 79)** |
| | Control | 5682 (24) | 18 490 | | |
| BNT162b2 (Pfizer-BioNTech) | Case | 179 (7) | 2569 | 73 (68, 77) | **74 (70, 78)** |
| | Control | 4772 (21) | 18 490 | | |
| mRNA-1273 (Moderna) | Case | 23 (<1) | 2569 | 82 (72, 88) | **82 (73, 89)** |
| | Control | 910 (5) | 18 490 | | |
| ChAdOx1 vaccines | Case | 87 (3) | 2569 | 58 (48, 66) | **63 (53, 71)** |
| | Control | 1492 (7) | 18 490 | | |

[a] VE estimates adjusted for gender (men, women); epidemiological week (14, 15, 16, 17, 18, 19, 20); and health authority (HA) (Fraser HA, Interior HA, Northern HA, Vancouver Coastal HA, Vancouver Island HA).
[b] VE estimates adjusted for age (50-59 years, 60-69 years); epidemiological week (14, 15, 16, 17, 18, 19, 20); and health authority (HA) (Fraser HA, Interior HA, Northern HA, Vancouver Coastal HA, Vancouver Island HA).

**Supplementary Table 7.** Single-dose vaccine effectiveness (≥21 days since vaccination) against hospitalization, overall and by variant of concern, adults 50-69 years, British Columbia, Canada

| Analysis | Case status | Sample sizes | | Vaccine effectiveness (VE) (95% CI) | |
|---|---|---|---|---|---|
| | | Vaccinated N (%) | Unvaccinated N | Crude | **Adjusted** |
| **Overall, weeks 14-20[a]** | | | | | |
| All vaccines | Case | 35 (6) | 547 | 82 (75,87) | **86 81,91)** |
| | Control | 12586 (26%) | 35358 | | |
| mRNA vaccines | Case | 32 (6) | 547 | 78 (69, 85) | **83 (76, 89)** |
| | Control | 9592 (21) | 35358 | | |
| BNT162b2 (Pfizer-BioNTech) | Case | 27 (5) | 547 | 78 (68, 85) | **83 (75, 89)** |
| | Control | 7959 (18) | 35358 | | |
| mRNA-1273 (Moderna) | Case | 5 (1) | 547 | 80 (52, 92) | **85 (63, 94)** |
| | Control | 1633 (4) | 35358 | | |
| ChAdOx1 vaccines | Case | 3 (1) | 547 | 94 (80, 98) | **96 (86, 99)** |
| | Control | 2994 (8) | 35358 | | |
| **Overall, weeks 17-20[b,c]** | | | | | |
| All vaccines | Case | 28 (12) | 215 | 84 (76, 89) | **87 (81, 91)** |
| | Control | 9871 (45) | 12194 | | |
| mRNA vaccines | Case | 25 (10) | 215 | 80 (70, 87) | **84 (75, 90)** |
| | Control | 7122 (37) | 12194 | | |
| BNT162b2 (Pfizer-BioNTech) | Case | 20 (9.0) | 215 | 80 (69, 88) | **84 (75, 90)** |
| | Control | 5802 (32) | 12194 | | |
| mRNA-1273 (Moderna) | Case | 5 (2) | 215 | 79 (48, 91) | **83 (58, 93)** |
| | Control | 1320 (10) | 12194 | | |
| ChAdOx1 vaccines | Case | 3 (1) | 215 | 94 (81, 98) | **95 (85, 99)** |
| | Control | 2749 (18) | 12194 | | |
| **Alpha variant of concern, weeks 17-20[b]** | | | | | |
| All vaccines | Case | 11 (12) | 80 | 83 (68, 91) | **85 (72, 92)** |
| | Control | 9871 (45) | 12194 | | |
| mRNA vaccines | Case | 11 (12) | 80 | 76 (56. 87) | **79 (60, 89)** |
| | Control | 7122 (37) | 12194 | | |
| BNT162b2 (Pfizer-BioNTech) | Case | 9 (10) | 80 | 76 (53, 88) | **79 (58, 90)** |
| | Control | 5802 (32) | 12194 | | |
| mRNA-1273 (Moderna) | Case | 2 (2) | 80 | 77 (6, 94) | **80 (17, 95)** |
| | Control | 1320 (10) | 12194 | | |
| ChAdOx1 vaccines | Case | 0 | 80 | NE | **NE** |
| | Control | 2749 (18) | 12194 | | |
| **Gamma variant of concern, weeks 17-20[b]** | | | | | |
| All vaccines | Case | 11 (10) | 94 | 86 (73, 92) | **89 (78, 94)** |
| | Control | 9871 (45) | 12194 | | |
| mRNA vaccines | Case | 8 (8) | 94 | 85 (70, 93) | **88 (76, 94)** |
| | Control | 7122 (37) | 12194 | | |
| BNT162b2 (Pfizer-BioNTech) | Case | 7 (7) | 94 | 84 (66, 93) | **88 (74, 95)** |
| | Control | 5802 (32) | 12194 | | |
| mRNA-1273 (Moderna) | Case | 1 (1) | 94 | 90 (29, 99) | **91 (36, 99)** |
| | Control | 1320 (10) | 12194 | | |
| ChAdOx1 vaccines | Case | 3 (3) | 94 | 86 (55, 96) | **90 (67, 97)** |
| | Control | 2749 (18) | 12194 | | |

[a] All VE estimates adjusted for age group (50-59 years; 60-69 years); gender (men, women); epidemiological week (14, 15, 16, 17, 18, 19, 20); and health authority (HA) (Fraser HA, Interior HA, Northern HA, Vancouver Coastal HA, Vancouver Island HA).
[b] All VE estimates adjusted for age group (50-59 years; 60-69 years); gender (men, women); epidemiological week (17, 18, 19, 20); and health authority (HA) (Fraser HA, Interior HA, Northern HA, Vancouver Coastal HA, Vancouver Island HA).
[c] Analysis of VE against variants of concern (VOC) was restricted to weeks 17-20 when full genetic characterization of case viruses was undertaken. VE against hospitalization due to Delta lacked sufficient sample size for reliable estimation, notably by vaccine type. VE against hospitalization due to non-VOC lacked sufficient sample size for reliable estimation overall or by vaccine type.

**Supplementary Table 8.** Single-dose vaccine effectiveness (≥21 days since vaccination) by variant of concern (VOC), adults 50-69 years, epi-weeks 17-20, British Columbia, Canada

| Vaccines | Case Status | Any Case, epi-weeks 17-20 | | | | Non-VOC [a,b] | | | | Alpha (B.1.1.7) [a] | | | | Gamma (P.1) [a,c] | | | | Delta (B.1.617.2) [a] | | | |
|---|---|---|---|---|---|---|---|---|---|---|---|---|---|---|---|---|---|---|---|---|---|
| | | Vac N (%) | Unvac N | Crude VE % (95%CI) | **Adj[d] VE % (95%CI)** | Vac N (%) | Unvac N | Crude VE % (95%CI) | **Adj[d] VE % (95%CI)** | Vac N (%) | Unvac N | Crude VE % (95%CI) | **Adj[d] VE % (95%CI)** | Vac N (%) | Unvac N | Crude VE % (95%CI) | **Adj[d] VE % (95%CI)** | Vac N (%) | Unvac N | Crude VE % (95%CI) | **Adj[d] VE % (95%CI)** |
| All | Case | 398 (19) | 1730 | 72 (68, 75) | **74 (70, 77)** | 13 (9) | 139 | 88 (80, 93) | **86 (75, 92)** | 175 (18) | 774 | 72 (67, 76) | **74 (69, 78)** | 140 (19) | 591 | 71 (65, 76) | **74 (68, 78)** | 17 (25) | 50 | 58 (27, 76) | **74 (53, 85)** |
| | Control | 9871 (45) | 12 194 | | | 9871 (45) | 12 194 | | | 9871 (45) | 12 194 | | | 9871 (45) | 12 194 | | | 9871 (45) | 12 194 | | |
| mRNA | Case | 238 (12) | 1730 | 76 (73, 79) | **78 (75, 81)** | 11 (7) | 139 | 86 (75, 93) | **84 (71, 92)** | 106 (12) | 774 | 77 (71, 81) | **78 (73, 82)** | 73 (11) | 591 | 79 (73, 83) | **80 (74, 85)** | 11 (18) | 50 | 62 (28, 80) | **74 (47, 87)** |
| | Control | 7122 (37) | 12 194 | | | 7122 (37) | 12 194 | | | 7122 (37) | 12 194 | | | 7122 (37) | 12 194 | | | 7122 (37) | 12 194 | | |
| BNT162b2 (Pfizer-BioNTech) | Case | 203 (2) | 1730 | 75 (71, 79) | **77 (73, 80)** | 8 (5) | 139 | 88 (75, 94) | **86 (71, 93)** | 93 (11) | 774 | 75 (69, 80) | **77 (71, 81)** | 64 (10) | 591 | 77 (70, 82) | **79 (73, 84)** | 9 (15) | 50 | 62 (23, 81) | **74 (45, 88)** |
| | Control | 5802 (32) | 12 194 | | | 5802 (32) | 12 194 | | | 5802 (32) | 12 194 | | | 5802 (32) | 12 194 | | | 5802 (32) | 12 194 | | |
| mRNA-1273 (Moderna) | Case | 35 (2) | 1730 | 81 (74, 87) | **82 (75, 87)** | 3 (2) | 139 | 80 (37, 94) | **81 (39, 94)** | 13 (2) | 774 | 84 (73, 91) | **85 (74, 92)** | 9 (2) | 591 | 86 (73, 93) | **85 (71, 92)** | 2 (4) | 50 | 63 (-52, 91) | **73 (-14, 94)** |
| | Control | 1320 (10) | 12 194 | | | 1320 (10) | 12 194 | | | 1320 (10) | 12 194 | | | 1320 (10) | 12 194 | | | 1320 (10) | 12 194 | | |
| ChAdOx1 | Case | 160 (8) | 1730 | 59 (52, 65) | **65 (59, 71)** | 2 (1) | 139 | 94 (74, 98) | **92 (66, 98)** | 69 (8) | 774 | 60 (49, 69) | **66 (57, 74)** | 67 (10) | 591 | 50 (35, 61) | **60 (48, 69)** | 6 (11) | 50 | 47 (-24, 77) | **73 (35, 88)** |
| | Control | 2749 (18) | 12 194 | | | 2749 (18) | 12 194 | | | 2749 (18) | 12 194 | | | 2749 (18) | 12 194 | | | 2749 (18) | 12 194 | | |

DSV = Days since vaccination, the interval in days between the date of receipt of the first mRNA dose and the date of specimen collection
Vac = vaccinated; Unvac = Unvaccinated; VOC = variant of concern; VE = vaccine effectiveness; 95% CI = 95% confidence interval; Adj = Adjusted

[a] Defined as in Supplementary Material 1.
[b] Non-VOC estimates displayed here include variants of interest (VOI); however, excluding VOI, non-VOC estimates were similar for vaccines overall in the crude (92%; 95% CI 81, 96) and adjusted (90%; 95% CI 77, 96) [based on 96 cases: 6 (6%) vaccinated]; mRNA vaccines combined in the crude (89%; 95% CI 74, 95) and adjusted (87%; 95% CI 71, 95) [based on 96 cases: 6 (6%) vaccinated]; BNT162b2 in the crude (91%; 95% CI 75, 97) and adjusted (89%; 95% CI 69, 96)[based on 94 cases: 4 (4%) vaccinated]; and mRNA-1273 in the crude (79%; 95% CI 17, 95) and adjusted (85%; 95% CI 36, 96) [based on 92 cases: 2 (2%) vaccinated] but with insufficient sample size for this estimation for ChAdOx1 vaccines.
[c] Gamma (P.1) estimates displayed here are identified by whole genome sequencing and/or are 501Y, 484K and K417T positive, excluding specimens missing information for the 417 single nucleotide polymorphism (SNP). Including the additional specimens missing the 417 SNP, VE estimates are similar for vaccines overall in the crude (70%; 95% CI: 64, 75) and adjusted (74%; 95% CI: 68, 78) [based on 765 cases: 148 (19%) vaccinated]; mRNA vaccines combined in the crude (78%; 95% CI: 73, 83) and adjusted (80%; 95% CI: 74, 84) [based on 695 cases: 78 (11%) vaccinated]; BNT162b2 in the crude (77%; 95% CI 70, 82) and adjusted (79%; 95% CI 72, 84)[based on 686 cases: 69 (10%) vaccinated]; and mRNA-1273 in the crude (87%; 95% CI 74, 93) and adjusted (86%; 95% CI 73, 93) [based on 626 cases: 9 (1%) vaccinated] and for ChAdOx1 vaccines in the crude (50%; 95% CI: 35, 61) and adjusted (60%; 95% CI: 48, 69) [based on 687 cases: 70 (10%) vaccinated].
[d] VE estimates adjusted for age group (50-59, 60-69 years); gender (men, women); epidemiological week (14, 15, 16, 17, 18, 19, 20); and health authority (HA) (Fraser HA, Interior HA, Northern HA, Vancouver Coastal HA, Vancouver Island HA).
